# Topic modelling with ICD10-informed priors identifies novel genetic loci associated with multimorbidities in UK Biobank

**DOI:** 10.1101/2022.10.28.22281623

**Authors:** Yidong Zhang, Xilin Jiang, Alexander J Mentzer, Gil McVean, Gerton Lunter

## Abstract

Studies of disease incidence have identified thousands of genetic loci associated with complex traits. However, many diseases occur in combinations that can point to systemic dysregulation of underlying processes that affect multiple traits. We have developed a data-driven method for identifying such multimorbidities from routine healthcare data that combines topic modelling through Bayesian binary non-negative matrix factorization with an informative prior derived from the hierarchical ICD10 coding system. Through simulation we show that the method, treeLFA, typically outperforms both Latent Dirichlet Allocation (LDA) and topic modelling with uninformative priors in terms of inference accuracy and generalisation to test data, and is robust to moderate deviation between the prior and reality. By applying treeLFA to data from UK Biobank we identify a range of multimorbidity clusters in the form of disease topics ranging from well-established combinations relating to metabolic syndrome, arthropathies and cancers, to other less well-known ones, and a disease-free topic. Through genetic association analysis of inferred topic weights (topic-GWAS) and single diseases we find that topic-GWAS typically finds a much smaller, but only partially-overlapping, set of variants compared to GWAS of constituent disease codes. We validate the genetic loci (only) associated with topics through a range of approaches. Particularly, with the construction of PRS for topics, we find that compared to LDA, treeLFA achieves better prediction performance on independent test data. Overall, our findings indicate that topic models are well suited to characterising multimorbidity patterns, and different topic models have their own unique strengths. Moreover, genetic analysis of multimorbidity patterns can provide insight into the aetiology of complex traits that cannot be determined from the analysis of constituent traits alone.

## Introduction

Multimorbidity, defined as the co-existence of multiple chronic conditions, is a major challenge for modern healthcare systems. Its prevalence has increased because of a worldwide increase in life expectancy ^1–3^, and it is associated with substantially lower quality of life ^3, 4^, worse clinical outcomes ^3^, and increased healthcare expenditure ^5^. The management of multimorbidity is challenging given that most guidelines and research are still targeted at single diseases. As a result, the negative impact of multimorbidity is often greater than the additive effects of individual diseases ^6^.

Several common multimorbidity patterns, such as a cluster composed of cardiovascular and mental health disorders, and a musculoskeletal disease cluster, have been identified from literature reviews ^3, 7^. In recent years, the widespread adoption of electronic health records (EHR) has enabled the systematic study of multimorbidity, and a variety of approaches have been employed for this purpose, including factor analysis ^8^, clustering ^9^, graph or network based methods ^10, 11^, and statistical models such as latent class analysis ^12–14^. These approaches have both validated the previously identified multimorbidity patterns ^12, 14, 15^ and, through the inclusion of a wider range of diseases, identified additional multimorbidity patterns ^14, 16^. In addition, downstream analyses enabled by these approaches have helped to identify the clinical events and outcomes associated with specific multimorbidity patterns ^13, 17^, which may provide insights about early intervention and risk stratification for patients.

The existence of common multimorbidity patterns raises the question of their etiology. One way to approach this question is to analyse multimorbidity patterns together with appropriateomics data to determine the biological pathways involved. These analyses have been made possible with the establishment of biobanks linking individuals’ biological samples and genetic information to their EHR ^18–20^. A recent study investigating genome-wide association studies (GWAS) of 439 common diseases recorded in UK biobank (UKB) hospital inpatient data found that 46% multimorbidity disease pairs have evidence for shared genetics ^11^, suggesting that this may be a fruitful approach.

Intrinsic to the study of multimorbidity is the joint analysis of multiple disease phenotypes. To enable this, various multi-trait GWAS methods have been developed which promise to better exploit the deep phenotype data available for individuals in biobanks. These methods can be subdivided based on their analytical approaches. Univariate methods combine signals from single-trait GWAS ^21–27^, while multivariate methods offer improved power by directly modelling the individual level genotype and phenotype data ^28–33^. Several of these methods use transformations such as principal components analysis (PCA) on the original traits before association analysis so that very large numbers of traits can be handled. Topic models such as Latent Dirichlet Allocation (LDA) and Non-negative Matrix Factorization (NMF) were dimension reduction algorithms developed to model word occurrence in text documents, and have subsequently found application in biological studies to extract complex patterns from high-dimensional data ^34–36^. They can be used to find multimorbidity clusters from diagnostic data, by viewing individuals as “documents” and diseases as “words”. The topics learnt by these models then are mathematical representations of groups of diseases that tend to co-occur within the same individual ^37^. Earlier studies have shown that joining single diseases into topics increases statistical power for genetic discovery, and helps to disentangle the pleiotropic effects of several known genetic loci ^38–40^.

Despite these advances, existing methods all have limitations. First, diagnostic data is often binary in nature, with zeros and ones representing the absence and presence of diseases, yet topic models like LDA and NMF were designed for count data, while algorithms designed for binary data ^41^ have not found wide application in biomedical studies. Second, biobank data is often sparse and, particularly for less common diseases, inclusion of prior domain knowledge may well improve results. Domain knowledge has been used successfully in topic models ^42, 43^, and, for example, medical ontologies like the ICD-10 disease classification system could serve as prior for disease co-occurrence, as they encode the complex relationships of diseases as a hierarchical structure which is amenable to mathematical analysis ^44–46^. Third, while for statistical models, such as LDA, principled approaches exist for selecting the number of clusters and optimising other hyperparameters ^47–51^, this is often not true for other methods, and these choices can strongly impact the final results ^9, 52^. In addition, methods not based on statistical foundations typically lack estimates of uncertainty in the inferred clusters, which makes interpretation difficult.

Here, we develop and validate an analytic framework for the study of multimorbidity using topic models and multi-trait GWAS on biobank datasets. Central to our approach is “treeLFA” (latent factor allocation with a tree-structured prior), a statistical model to identify multimorbidity clusters of common diseases based on co-occurrence patterns and an informed prior derived from a tree-structured disease ontology. Applying treeLFA to Hospital Episode Statistics (HES) data extracted from UKB we gain insights about the relationships of diseases and their shared genetic components. We identify multimorbidity clusters in the form of disease “topics” and show that these agree with accepted medical understanding. Performing a series of GWAS on the quantitative traits defined by individuals’ weights for these topics (topic-GWAS), we show that the approach identifies novel loci that correlate in expected ways with several genomic annotations. We validate the topic-GWAS results using test data, and show that topic-GWAS can improve genetic risk prediction for multiple disorders, in particular immune disorders, and those for which currently few associated loci are known.

## Results

### Overview of treeLFA

treeLFA is a topic model designed to identify multimorbidity clusters from binary disease diagnosis data. We describe the model in terms of the associated generative process. To generate data, first topic vectors *ϕ_k_* containing disease probabilities, and a topic weight vector *θ_d_* for each individual *d* are sampled from a prior distribution. An individual’s disease probabilities are given by a mixture of different topic vectors, with the topic weights (*θ_d_*) acting as mixing proportions. The model can equivalently be defined by the likelihood for observations, which involves factoring a latent matrix of disease probabilities:

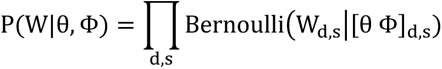

Here, *W* is the binary input matrix recording individuals’ diagnosed diseases, with rows representing individuals and columns representing disease codes; P(W_d,s_=1) is the probability of disease s being 1 (diagnosed) for individual d. *ϕ* is the topic-disease matrix (each row a topic and each column a disease), and *θ* the topic weights matrix (each row an individual and each column a topic). The occurrence of disease *s* for individual *d* is modelled with a Bernoulli distribution parameterized by the corresponding entry in the product of matrices *θ* and *ϕ*: [*θ ϕ*]*_d,s_*. This model differs from LDA in three ways. First, LDA samples diseases (or words) according to a multinomial distribution, so that diseases can occur multiple times, while treeLFA only allows presence or absence. Second, LDA conditions on the number of observed diseases, whereas for treeLFA the number of diseases is informative. Third, treeLFA uses an informative prior on topic vectors *ϕ_k_* guided by a tree-structured ontology such as ICD-10 (Figure 1 in the analytic note). This prior has the property that diseases that are closely related on the tree tend to have correlated probabilities. Inference on the treeLFA model is performed using partially collapsed Gibbs sampling ^53^, integrating out the topic weight variable. See the analytic note for more details, including on hyperparameter optimization.

**Figure 1.**
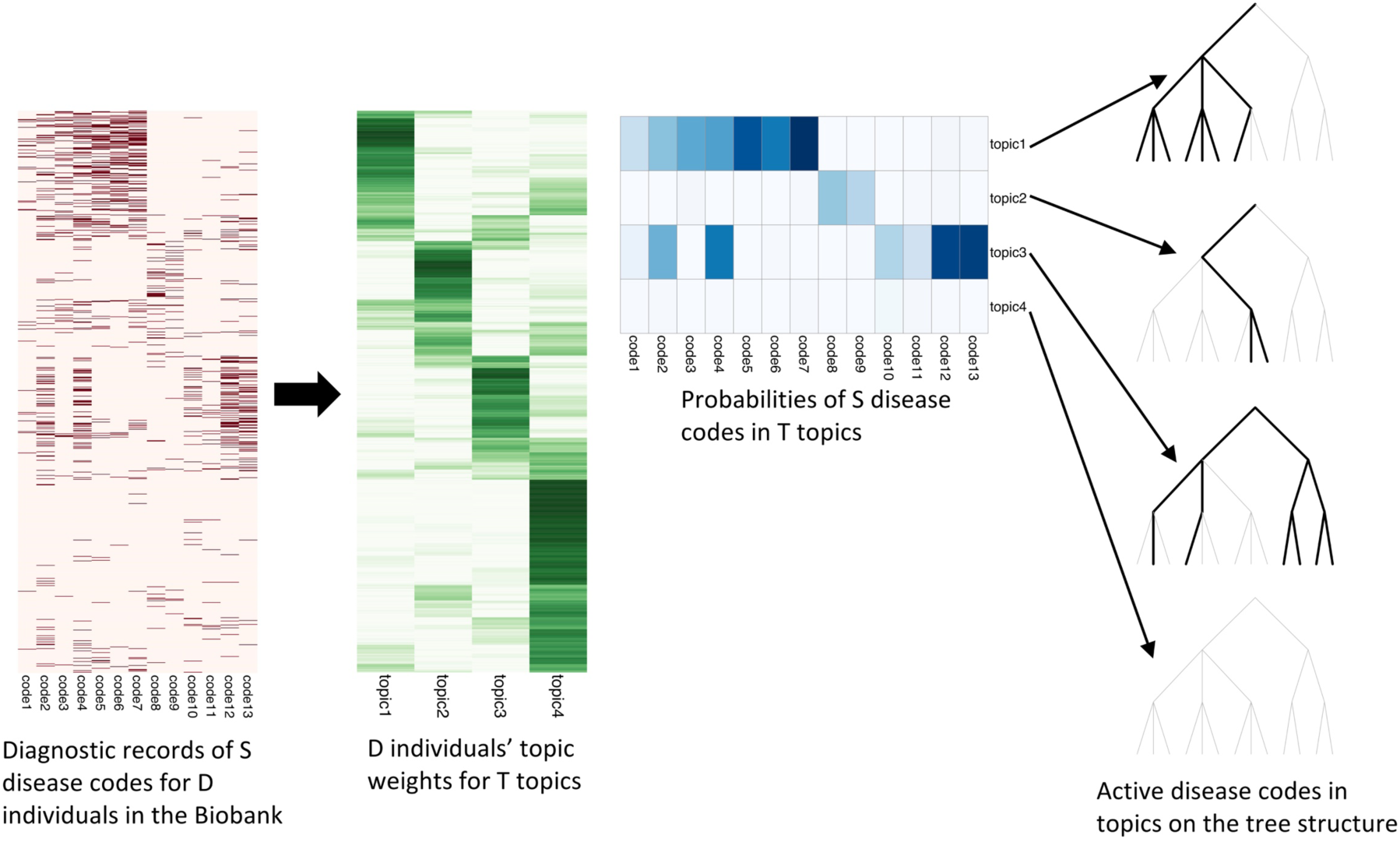
Schematic for topic modelling of the diagnosis data in UKB with treeLFA. The presence and absence of S disease codes for D individuals in the biobank is modelled with D×S Bernoulli distributions. The matrix of Bernoulli probabilities is factored into the product of a topic matrix and a topic weight matrix. Individuals’ weights for topics are modelled with categorical distributions with a Dirichlet prior. Each topic is composed of S probability variables with Beta priors to parameterize the Bernoulli distributions for disease codes. Disease codes can be either active or inactive in topics. Active disease codes have large probability while inactive ones have near zero probability. A prior for topics (specifies the likelihood of different disease codes to be active in topics) is constructed on the tree structure of disease codes specified by a medical ontology (such as the ICD-10 coding system). The path from the root node to active leaf nodes (corresponding to all active disease codes in a topic) are highlighted on a three-layered tree structure for 13 disease codes in 4 topics.

### Validation of treeLFA; Comparison with related topic models

We assessed treeLFA’s performance in a simulation experiment, comparing it to the same model but without an informative tree prior (flatLFA; Fig 2a), and to LDA. We designed the simulation to test the model with respect to the degree of multimorbidity in the data; the size of the data; and the correctness of the prior. The degree of multimorbidity was governed by α, the concentration parameter of the Dirichlet prior for the topic weight variable θ, with large values corresponding to the presence of several multimorbidity clusters in individuals, and small values resulting in individuals mainly presenting diseases from a single cluster. To test the influence of prior misspecification we used two sets of topics for simulation. In one set (“correct tree prior”) the active disease codes in topics were aligned with the tree structure of disease codes, resulting in a high likelihood under the prior, which specifies that child nodes on the tree tend to (though not exclusively) have the same activity as their parent nodes (Fig 2b,c). In the other setting (“incorrect tree prior”) the pattern of active disease codes in topics was possible but unlikely under the prior (Supplementary Fig 1a,b). For each set of topics we considered four combinations of D and α, resulting in eight parameter combinations in total (Supplementary Table 1). For each of these we generated 20 data sets. To evaluate the performance of treeLFA, flatLFA and LDA we used two metrics: the accuracy of the inferred topic (*Δϕ*, average absolute per-disease difference in probability between aligned true and inferred topics; see Methods for details), and R_pl_, the ratio of the average per-individual predictive test likelihood for treeLFA and flatLFA.

**Figure 2.**
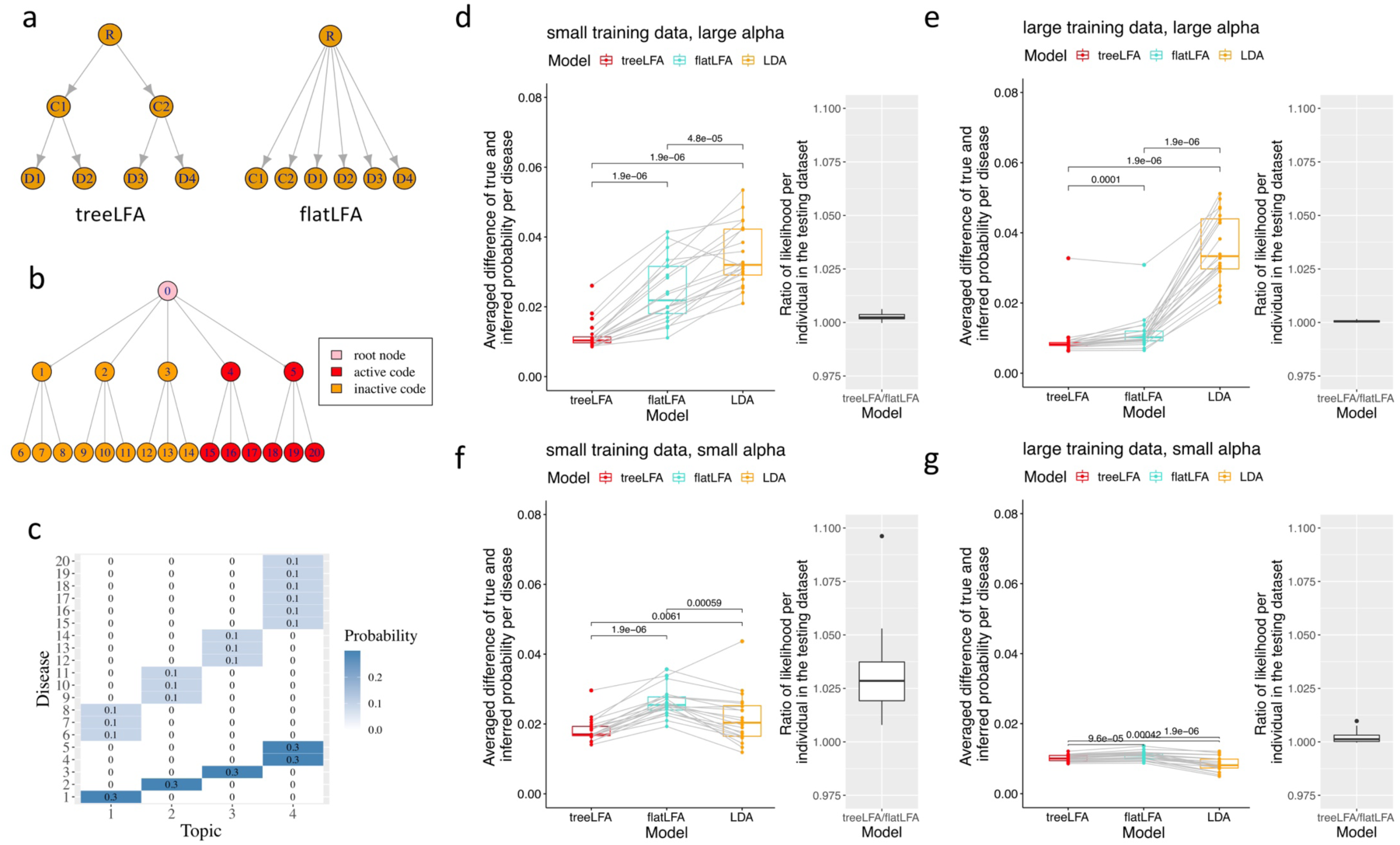
Comparison of three related topic models (treeLFA, flatLFA and LDA) on simulated datasets. a, The informative and non-informative tree structures used by treeLFA and flatLFA. b, The tree structure of the 20 disease codes used for simulation. Red nodes on the tree correspond to all active disease codes in Topic 4 used for simulation (in Figure 2c). c, The four topics used for simulation. The heatmap shows the probabilities of diseases in topics. Each row corresponds to a disease code, each column corresponds to a topic. Inactive disease codes in topics have zero probability. All active disease codes in each of the first three topics come from the same branch of the tree in Figure 2b. Active codes in the last topic come from the last two branches of the tree in Figure 2b. d-g, Comparison of three topic models on simulated datasets. The performance of three topic models (treeLFA, flatLFA and LDA) on four groups of simulated datasets are shown. The four groups of datasets were generated using the same topics (Figure 2c), and different values for D (number of individuals in the training dataset) and α (the concentration parameter of the Dirichlet prior for individuals’ topic weights). For each combination of D and α, 20 datasets (including both training and testing datasets) were simulated. Inference accuracy of topic models is evaluated using the averaged per disease difference between true and inferred probability of all diseases in all 4 topics (box plots). Each dot in a box plot is the result of one model on one dataset, and dots for different models on the same dataset are connected with grey lines. For treeLFA and flatLFA, the predictive likelihood on the testing datasets were calculated using topics inferred on the training data. Each dot in the point plot represents the treeLFA to flatLFA ratio of per individual averaged predictive likelihood for one dataset. d, Results on datasets simulated using D=2500 and α=1. e, Results on datasets simulated using D=5000 and α=1. f, Results on datasets simulated using D=300 and α=0.1. g, Results on datasets simulated using D=1000 and α=0.1.

Both metrics indicate that on data simulated using the correct tree prior, treeLFA performs better than flatLFA, which does not have the benefit of an informative prior (Figures 2d-g, Supplementary Table 2). This is most pronounced for small datasets with strong multimorbidity (Figure 2d; *Δϕ* 0.012±0.004 (treeLFA) and 0.025±0.010 (flatLFA); R_pl_ 1.003±0.002), while for larger datasets, the two models show similar performance and the prior has less influence (Figure 2e; *Δϕ* 0.009±0.006 (treeLFA) and 0.011±0.005 (flatLFA)). treeLFA outperforms LDA except for large data sets with weak multimorbidity (Figure 2g; *Δϕ* 0.0100±0.0009 (treeLFA) and 0.0084±0.0021 (LDA)) where both models gave accurate inference. For simulation using incorrect tree priors, flatLFA gave results comparable to simulation with correct priors, and the performance of flatLFA and treeLFA is similar across the four parameter combinations (Supplementary Figure 1), indicating that treeLFA is robust against prior misspecification. Overall, these results indicate that both treeLFA and flatLFA give accurate results when sufficient training data is available, even if treeLFA’s informative prior is inaccurate; but when the tree prior is correct, treeLFA performs better than flatLFA, particularly when training data is limited. Even in larger real-world data sets, low-frequency topics will have limited training data, hence this suggests that treeLFA could add power to the analysis of multimorbidity in biobank data.

### Topics of ICD-10 codes inferred from UK Biobank data

To investigate the properties of treeLFA on real-world data, we built an exploratory data set using the HES data in UKB from 502,413 individuals, consisting of the 100 most frequent codes from chapters 1-13 of the ICD-10 coding system (top-100 UKB dataset, Supplementary Table 3). We split these data randomly into training (80%) and testing (20%) datasets, and trained treeLFA with an initial K=11 topics (There is a discussion of the optimal number of topics below).

The inferred topics include an “empty” topic, in which all codes have near-zero probability of occurring (Figure 3a, Supplementary Table 4). Its associated entry in the optimal Dirichlet prior parameter *α* is very large (0.585) compared to that of other topics (0.016-0.06), indicating that the empty topic is frequently assigned to an individuals’ disease profile. The remaining topics all contain active codes. Most topics are sparse (8 topics contain fewer than 10 high-probability (>0.2) codes), but the model also infers dense topics, such as topics 8 and 10 which include 41 and 43 high-probability codes respectively. To assess whether codes tend to be specific to a topic, we normalised their probabilities across topics to make the largest probability 1. We found that, in general, codes are typically specific to topics: most (87/100) are active (normalised probability>0.5) in 3 or fewer topics. However, some codes are active in many topics, such as I10 (essential hypertension, active in 6 topics) and C44 (other malignant neoplasms of skin, active in 8 topics) (Figure 3a), suggesting that they have both large prevalence and a large number of multimorbidity partners belonging to different disease clusters. The top disease codes (i.e. the five with the largest probabilities) in the 10 non-empty topics are consistent with known disease mechanisms (Figure 3b), and are most frequently drawn from two (6/10) of the ICD-10 chapters. Specifically, in Topic 5, codes E78 (disorders of lipoprotein metabolism and other lipidemias) and I10 (essential hypertension) are components of the metabolic syndrome ^54^, which is associated with increased risk for cardiovascular diseases (CVD) ^55^, an association supported by the other three inferred top codes for this topic (I20, I21 and I25, all heart diseases). Another example is Topic 11, whose top codes include four spondylopathy subtypes, while the remaining one is G55 (nerve root and plexus compressions), a common complication of intervertebral disk disorders.

**Figure 3.**
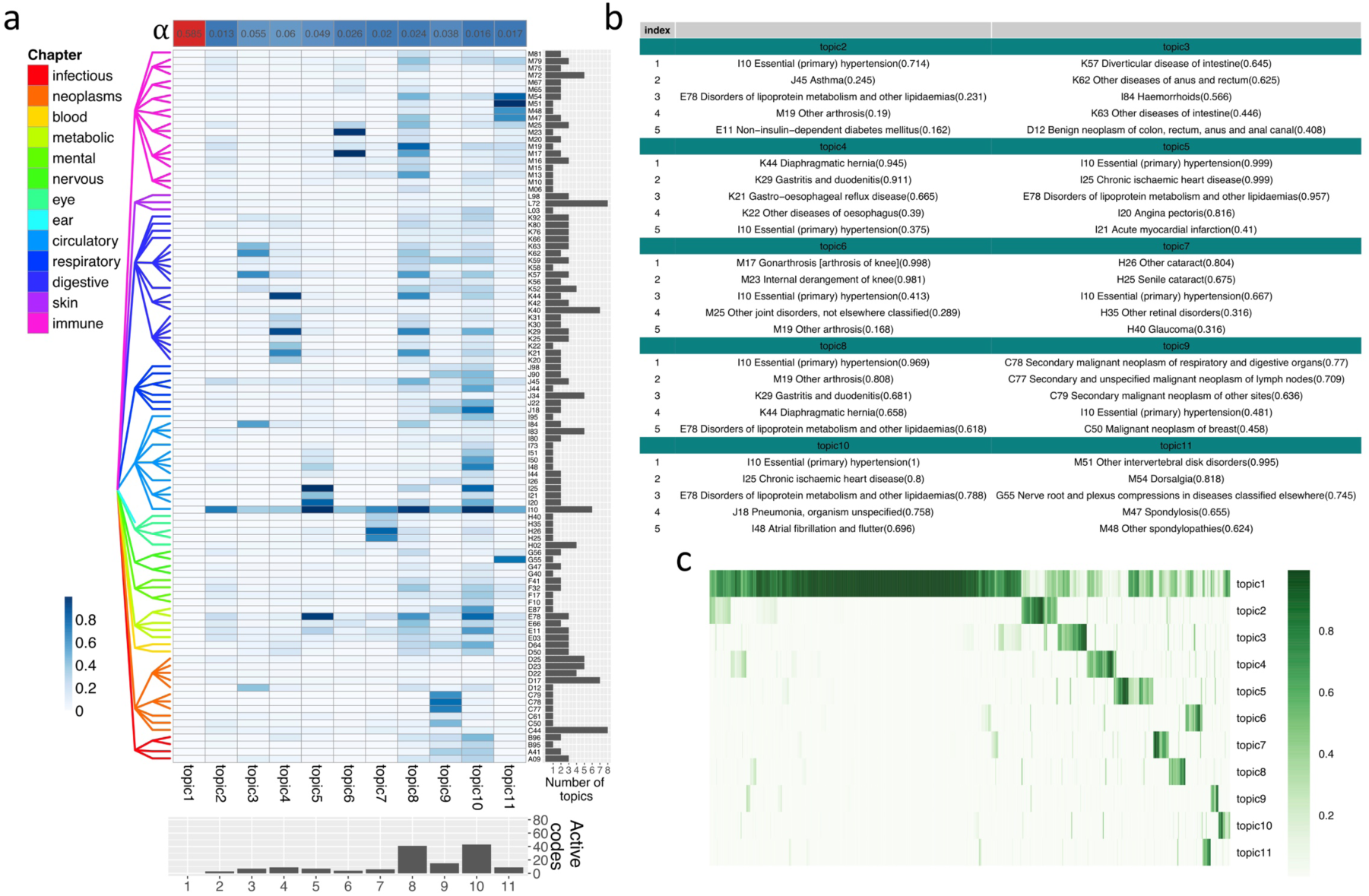
Inference results given by treeLFA on the top 100 UKB dataset. a, 11 topics inferred by treeLFA from the top-100 UKB dataset. The heatmap shows the probabilities of 100 ICD-10 codes in the 11 treeLFA topics, in which each row is an ICD-10 code and each column is a topic. Topics are arranged in a descending order of their corresponding entries in the optimised α vector (the single-row heatmap on the top). The tree structure for the 100 ICD-10 codes is shown to the left of the heatmap. Codes from different chapters of the ICD-10 coding system are colored differently. The barplot below the heatmap shows the numbers of ICD-10 codes with a probability of at least 0.2 in topics. The barplot on the right side of the heatmap shows the number of topics in which an ICD-10 code is active (with a normalised probability of at least 0.5). b, The top 5 codes with the largest probability in the 10 non-empty treeLFA topics (topics 2-11 in Figure 3a). Numbers in the brackets show the probabilities of disease codes in topics. c, Inferred weights for the 11 topics for 2000 random individuals. Each row in the heatmap is a topic, and each column is an individual.

In addition to defining topic vectors, the model also infers individuals’ weights for all topics (shown for 2,000 individuals in Figure 3c, Supplementary Table 5). As expected, most individuals have substantial weight for the empty topic, and this weight was strongly and negatively associated (Pearson correlation: -0.853) with the total number of diagnoses. Individuals that were not diagnosed with any of the top-100 ICD-10 codes (629/2,000) have a weight near 1 for the empty topic, while the majority of other individuals (1056/1371) have large weight (>0.1) for less than two disease (non-empty) topic, as expected from the sparsity of the data.

To compare the performance of treeLFA with flatLFA and LDA, we used the same input data to train the flatLFA model with 11 topics and the LDA model with 10 topics (no empty topic would be inferred by LDA, so it was trained with one fewer topic). Topics inferred by treeLFA and flatLFA were almost identical (Supplementary Figure 2a), indicating that the input data was large enough to make the impact of the informative prior minimal. Most topics inferred by LDA also had a high level of similarity to the non-empty topics inferred by treeLFA, except for two topics, for which the cosine similarities were 0.685 and 0.853 (Supplementary Figure 2b). Overall, these results indicate that the three topic models captured the same multimorbidity pattern from the top-100 UKB dataset.

### GWAS on topic weights

We next investigated whether the quantitative traits defined by the topic weights can be used to identify genetic variants that are associated with an individual’s risk for developing multimorbidities represented by the topics. We performed GWAS on individuals’ weights for the 11 topics inferred by treeLFA (topic-GWAS), and identified associations that reached genome-wide significance (p<5×10^-8^; non-lead SNPs with r^2^>0.1 were removed). For comparison, we also performed standard binary GWAS for the 100 ICD-10 codes and the 296 Phecodes mapped from these ICD-10 codes (see Methods for details).

We found 128 independent loci associated with at least one of the 11 topics, while 812 independent loci were associated with at least one of the 100 ICD-10 codes; 82 loci were shared between the sets (Figure 4a). Phecode GWAS showed similar patterns (Supplementary Figure 3a,b). Breaking this down by topic, we find that unique loci found by topic-GWAS were highly non-randomly distributed (Figure 4b, Supplementary Table 6). Most unique loci were associated with the empty topic (20/36), followed by Topic 8 (17/28) which contains a large number (41) of high probability codes (>0.2) from Chapter 11 (Diseases of the digestive system, 12 codes) and 13 (Diseases of the musculoskeletal system and connective tissue, 13 codes). In contrast, four sparse topics showed no unique loci. Topics 5 (metabolic and heart diseases) and 6 (joint diseases) had many associated loci (50 and 17 respectively) and also a substantial number of unique loci (5 and 7 respectively), suggesting that active codes in these topics include shared genetic components. The identification of novel loci indicates that topic-GWAS provides additional power for discovery. For example, Figure 4c (Supplementary Table 7) compares P-values of lead SNPs from the topic-GWAS for Topic 5, and P-values for association of the same loci with the top five active codes in Topic 5 (E78, I10, I20, I21, I25) from the single code GWAS. For most topic-associated lead SNPs, P-values given by topic-GWAS are smaller than those given by the corresponding single code GWAS, indicating increased power for these examples. This also explains some loci uniquely found by topic-GWAS, including some loci that show single-code P-values well below genome-wide significance (see Figure 4d for two example loci). Despite the limited numbers of topic-associated loci, the genomic control inflation factor and LD score regression (LDSC) indicate that most topics are in fact highly polygenic traits, with the exception of the empty topic and Topic 8, for which LDSC analysis suggests that uncontrolled confounding factors exist (Supplementary Table 8; age, sex and the first 10 PCs were controlled for in topic-GWAS).

**Figure 4.**
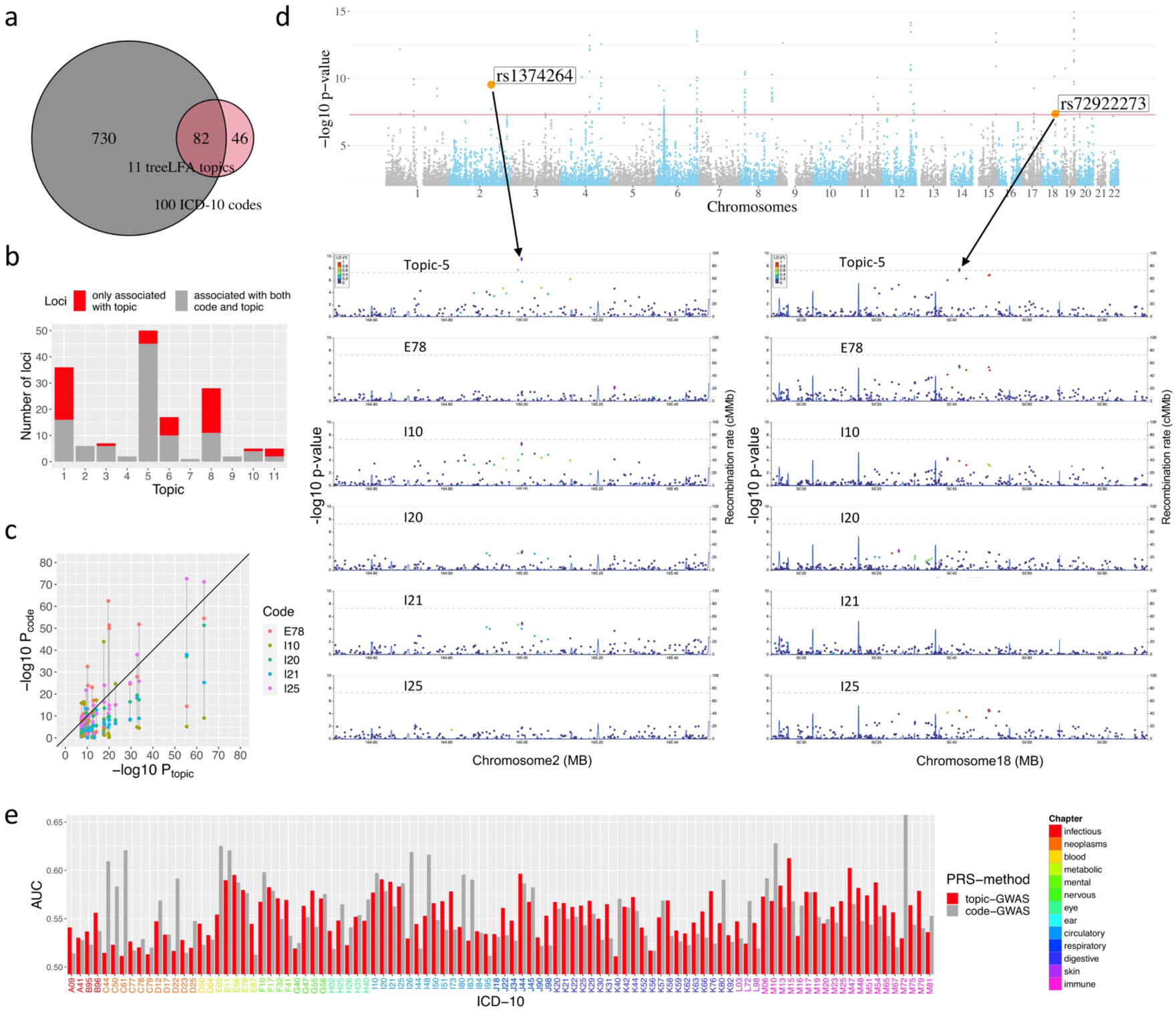
topic-GWAS result for the 11 topics inferred by treeLFA. a, The total numbers of significant loci found by topic-GWAS for the 11 treeLFA topics and single code GWAS for the 100 ICD-10 codes, and the overlap of these two sets of loci. b, The numbers of significant loci found by both single code/topic-GWAS, and the numbers of loci only found by topic-GWAS for the 11 treeLFA topics. c, Comparison of P-values given by topic-GWAS for all lead SNPs for Topic 5 and P-values for the same SNPs given by single code GWAS for the top 5 active codes (E78, I10, I20, I21, I25) in Topic 5. d, The Manhattan plot for Topic 5, and the regional Manhattan plots for single code/topic-GWAS results for two example lead SNPs of Topic 5. e, Comparison of two types of PRS for the 100 ICD-10 codes. One type of PRS is directly constructed using the single code GWAS results. Another type of PRS for ICD-10 codes is constructed as the sum of PRS for topics weighted by the probabilities of an ICD-10 code in all topics. The AUC of these two types of PRS on the test dataset are plotted.

We next asked whether topic-GWAS simply identify associations with disease groups (categories) represented by internal nodes of the ICD-10 or the Phecode ontology tree, which correspond to expert-led disease clusters and provide a useful contrast to our data-driven multimorbidity clusters. To answer this we performed GWAS on groups of ICD-10 codes or Phecodes corresponding to internal nodes in the respective classification systems. We found 634 loci associated with the 68 internal ICD-10 codes and 296 loci associated with the 136 internal Phecodes. Of the 128 topic-associated loci, 41 were not associated with any of the internal or terminal ICD-10 codes; and for Phecodes the corresponding number was 56 (Supplementary Figure 3c-f). This indicates that topic modelling provides insights into the relationships of diseases beyond those provided by expert-driven disease groupings encoded in ontologies. For example, Topic-8 has the majority of its active codes coming from Chapter 11 (Diseases of the digestive system) and 13 (Diseases of the musculoskeletal system and connective tissue), and a similar multimorbidity cluster was also identified by a recent study on UK Biobank ^11^. Interestingly, this cluster has many unique loci found by topic-GWAS, possibly indicating that these two categories of diseases share some underlying biology.

We then compared the topic-GWAS results for topics inferred by treeLFA, flatLFA and LDA. As expected from the similarity of topics inferred by treeLFA and flatLFA, similar numbers of associated loci were identified (128 and 126; Supplementary Figure 4a), compared to LDA, which identified many fewer (65; Supplementary Figure 4b), of which 44 overlap with the treeLFA loci. This difference in numbers of associated loci is mostly due to treeLFA’s empty topic (associated with 36 loci), which is not identified by LDA, and also due to differences in the dense Topic 8 (treeLFA, 28 loci; LDA, 4 loci), and topics 5 and 6 (Supplementary Figure 4c). One reason for the relatively poor performance of LDA may be that LDA-derived topic weights are negatively correlated with each other, as they must sum to one, while treeLDA’s topic weights are only negatively correlated with the empty topic weight, but are otherwise almost independent of each other (Supplementary Figure 5).

### Validation of topic-GWAS results

To exclude the possibility that the unique topic-GWAS associations were driven largely by technical biases or population stratification, we validated the results in three ways. First we considered overlap with previously identified loci reported in the GWAS catalogue ^56^. We find that 114/128 (89.1%) of all topic-associated loci and 36/46 (78.3%) of unique associations have records in the GWAS catalogue, and this overlap is consistent across topics (Supplementary Figure 6a). Second, we looked at enrichment of topic-associated loci in functional genomic regions. To do this we defined three groups of SNPs, including lead SNPs for all loci associated with ICD-10 codes, a random selection of 10,000 GWAS tag SNPs (controls) and topic-associated lead SNPs that were not found by single code GWAS, and then compared the proportions of them that have different functional properties (two-proportion Z-test, adjusted P-value<0.05, Bonferroni correction). We find that compared to random SNPs, a significantly larger proportion of topic-associated SNPs are in genomic regions with strong transcription activity (using chromHMM-predicted chromatin states as proxy ^57^). In addition, the proportions of SNPs that are QTLs and have chromatin interactions (CI) in at least one tissue in the first and third groups are similar (0.83 and 0.77 are eQTL, 0.90 and 0.98 have CI), and larger than the corresponding proportions in the control group (0.50 and 0.65 for eQTL and CI), indicating that loci associated with single codes and topics have comparable functional properties which are different from those for controls. (Supplementary Figure 6b-d).

Third, we made use of the test data to validate topic-GWAS results. We reasoned that if topics and their associated loci represent true biological processes, then topic-GWAS results should enable us to predict the risks of individual diseases with an accuracy comparable to that achieved using single code GWAS. To do this we first constructed PRS for topic weights using topic-GWAS results on training data. We found that they all show significant association with inferred topic weights on test data (Supplementary Table 9, see Methods for details). We then used these PRS for topics to construct PRS for the 100 ICD-10 codes, by adding individuals’ PRS for the ten disease topics weighted by the probability of the ICD-10 code of interest in each topic. For comparison, we also constructed PRS for all ICD-10 codes directly using the single-code GWAS results in the standard way. Each pair of PRS for an ICD-10 code was used to predict individuals’ corresponding diagnosed disease in the test data, and their performance was evaluated using the area under the receiver-operator curve (AUC) statistic. For 65 ICD-10 codes, topic-PRS AUCs are larger than single-code PRS AUC (Figure 4f, Supplementary Table 10), with increases ranging from 1% to 5%. This increase was seen most for ICD-10 codes from chapters 5 (Mental and behavioural disorders, 75% (3/4) showing increased AUC), 11 (Diseases of the digestive system, 86% (18/21)) and 13 (Diseases of the musculoskeletal system and connective tissue, 70% (14/20)). By contrast, single-code PRS performed well for codes that have a relatively large number of associated loci found by single code GWAS (>10 associated loci; 18/22 disease codes show larger AUC for single-code PRS than topic-PRS; Supplementary Figure 7a,b). Finally, to make an objective comparison of the topic-GWAS for treeLFA and LDA, we constructed PRS for ICD-10 codes from LDA’s topic- GWAS using the same approach, and found that in the majority of cases (99/100) the PRS based on treeLFA’s results have larger AUC (Supplementary Figure 7c), indicating treeLFA’s topic-GWAS is more informative. Taken together, the three complementary approaches indicate that topic-GWAS associations broadly represent true genetic associations with biological phenotypes.

### Inference and topic-GWAS results across models

Before applying treeLFA to a larger data set containing more diseases, we considered how to select the number of topics (K), a fundamental problem for topic models. We trained treeLFA models with different numbers of topics (K=2-20, 50, 100) on the top-100 data set, and found that for larger K topic vectors were frequently duplicated, therefore we performed clustering on the posterior samples of topics (see Methods for more details). The resulting topics always included an empty topic, and as K increased the topics tended to become more sparse, although some dense topics always remained (Figure 5a, Supplementary Table 11). As K increased, topics tended to split into sub-topics, which we visualised in a tree by connecting each topic to its most similar topic (measured by Pearson correlation of topic vectors) in the layer above, and we observed that the topics split in a stable way (Figure 5b, Supplementary Table 12). These observations indicate that topic-GWAS loci and associated effect sizes should also be stably identified. We verified this for many loci (examples in Supplementary Figure 8), and Figure 5b illustrates this for a single variant. On the top-100 UKB dataset, the number of distinct topics remaining after clustering is saturated at 25-30 topics (Supplementary Figure 9a). Similarly, the total number of topic-GWAS loci, the number of unique such loci, and the predictive likelihood on the test data all began to saturate beyond K=20 (Figure 5b; Supplementary Figure 9b). We do note that for models with K=50 or K=100, we infer several near-empty topics after clustering, which are unlikely to be stable multimorbidity patterns and are challenging to interpret. Taken together, these results indicate that selecting a sufficiently large value for K, combined with post-hoc clustering of topics, is a computationally efficient strategy for producing a stable and comprehensive set of topics.

**Figure 5.**
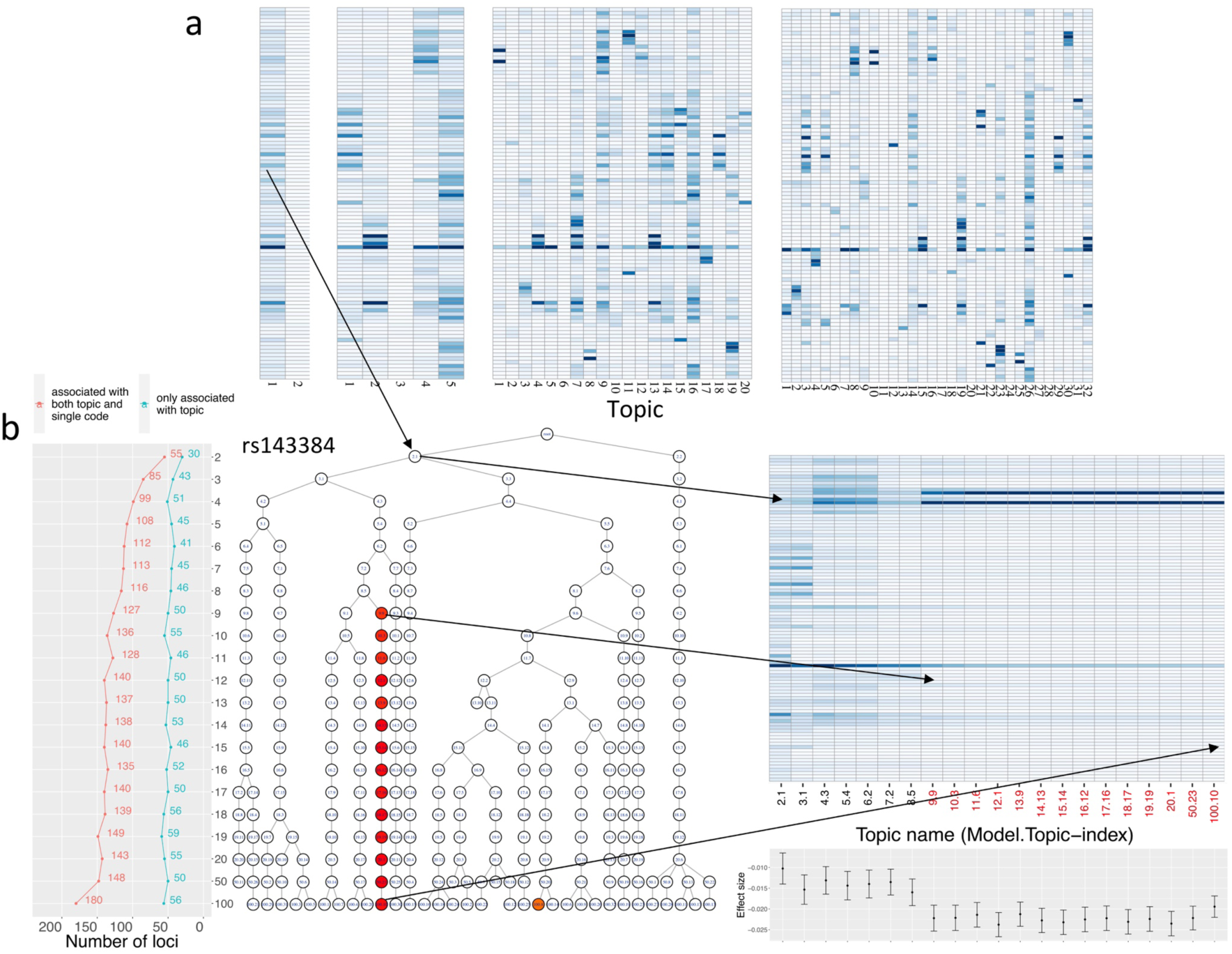
Inference and topic-GWAS results across treeLFA models set with different numbers of topics. a, Averaged inferred topics given by the treeLFA models set with 2, 5, 20 and 100 topics. For each treeLFA model, ten Gibbs chains were trained, and 50 posterior samples of topics were collected from each chain. Posterior samples of topics from different chains are mixed and clustered, and the mean topic vectors are then calculated for each cluster. b, Relationship of topics inferred by different models. All topics inferred by all treeLFA models are organised into a tree structure. Each node on the tree is a topic inferred by a model, and all nodes on the same layer (level) of the tree are all topics inferred by the same treeLFA model (set with a certain number of topics). Each topic in the tree is connected to its most similar topic (measured with the Pearson correlation) in the layer above. Topics associated with the SNP rs143384 are colored according to the –log10(P-value) for the SNP in the corresponding linear regression. Most of the associated topics are in the same branch of the tree, so all topics in this branch are plotted in the heatmap on the right side of the tree, with names of topics (model.topic-index) associated with rs143384 highlighted in red. In the barplot below the heatmap, effect sizes and standard errors of rs143384 given by topic-GWAS for the above topics are plotted. The line plot to the left of the tree shows the total numbers of topic-associated loci and the numbers of topic-associated loci that are not found by single code GWAS for different treeLFA models.

### Results on a larger UKB dataset

We next defined a larger data set consisting of the 436 ICD-10 codes from chapters 1-14 with a prevalence exceeding 0.001 in UKB (top-436 UKB dataset, Supplementary Table 13), and again randomly split this 80/20 into training and testing datasets. Training treeLFA/flatLFA models with 100 topics, we identified about 40 distinct topics after clustering of posterior samples. Therefore, we kept 40 topics for both models (for the convenience of an objective comparison of predictive likelihood), and collapsed the remaining near-empty topics into the empty topic.

Since the inference results (topics) given by different treeLFA/flatLFA chains were not exactly the same, we used the result given by the chain with the largest predictive likelihood on the test data for the downstream analyses. Among the 40 inferred topics, 29 were found by all three treeLFA chains, and five were found by two treeLFA chains, suggesting most topics were stably identified from the data (Figure 6a, Supplementary Table 14). The 40 topics again include several dense topics, with topics 1-5 including more than 40 active codes (having a normalised probability>0.5 in a topic). The set includes many sparse topics, with most including active codes enriched (Fisher exact test, adjusted P values<0.05, FDR corrected) for 1-2 ICD-10 chapters, and again a single empty topic (Figure 6a-b; Supplementary Table 14,15). The top active codes in topics (defined as having an unnormalized probability>0.3) are shown in Table 1, where topics are annotated based on the categories of these top active codes. For most topics, their top active codes represent similar diseases, such as diseases affecting the same physiological system or having the same pathological mechanism. Comparing topics identified by treeLFA and flatLFA, we found that 32 topics were identified by both models (cosine similarity>0.9; Supplementary Figure 10a), while the remaining topics have substantial differences. Overall, the predictive likelihood of treeLFA chains was better than that of flatLFA, and has a smaller range (Supplementary Figure 10b), indicating that the tree-based prior is indeed helpful in extracting meaningful patterns from the data.

**Figure 6.**
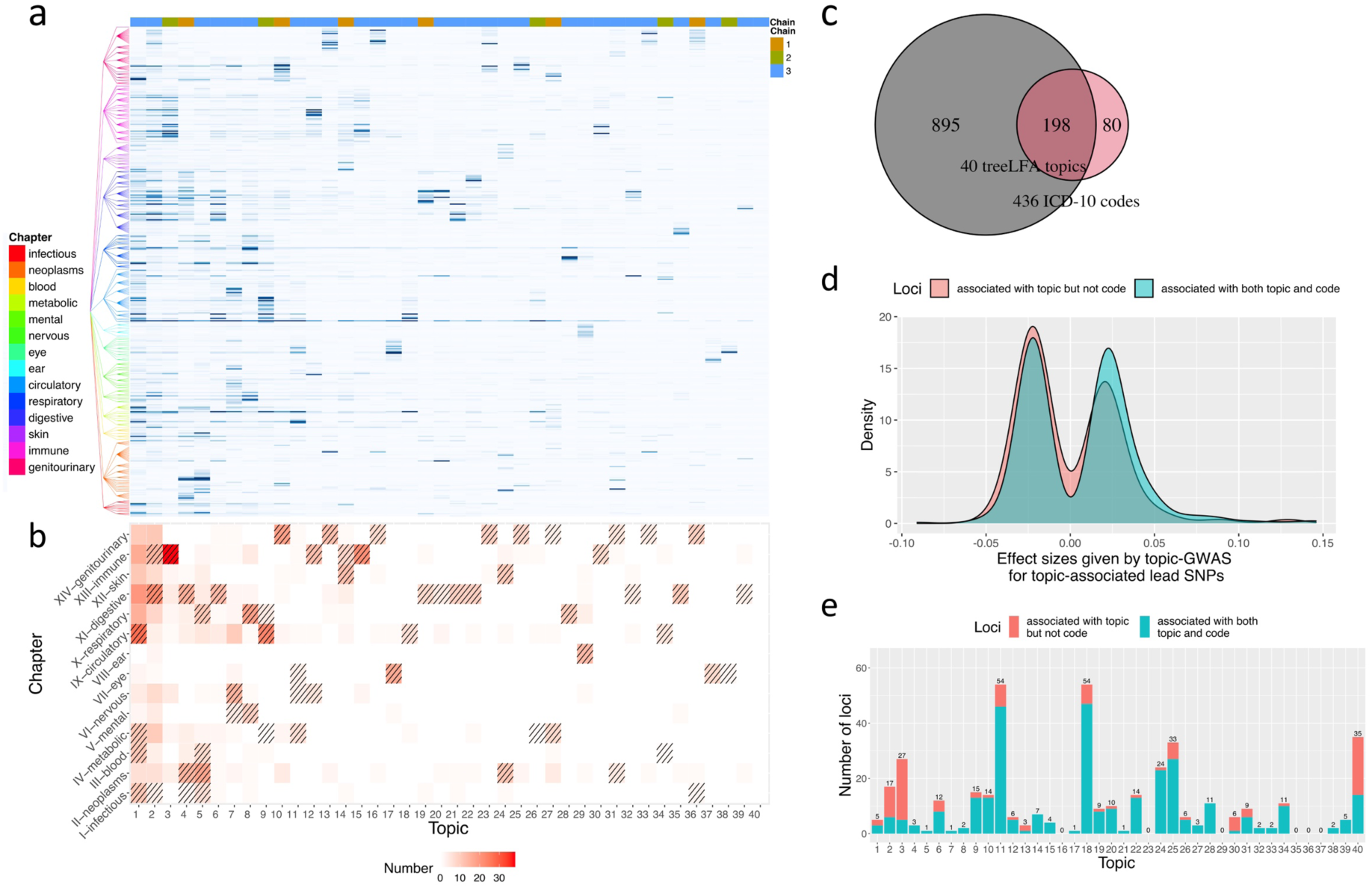
Inference and topic-GWAS results for the 40 topics inferred by treeLFA from the top-436 UKB dataset. a, The 40 topics inferred by the treeLFA model set with 100 topics. Topics are ordered according to their density (the sum of the probability of all codes in the topic). The tree structure of the 436 ICD-10 codes is plotted to the left of the heatmap. The colour bar on top shows for each topic the number of treeLFA chains which inferred it. b, The numbers of active codes (with a normalised probability of at least 0.5) in different topics coming from different ICD-10 chapters. Enriched chapters among active codes in topics are highlighted with shades in the cells (Fisher exact test, FDR<0.05). c, The total numbers of loci associated with the 40 treeLFA topics and the 436 ICD-10 codes, and the overlap of these two sets of loci. d, Distribution of effect sizes given by topic-GWAS for lead SNPs associated with only topic and lead SNPs associated with both topic and single code. e, The total numbers of loci associated with different topics (in the same order as topics in Figure 6a), and the numbers of topic-associated loci that are not found by single code GWAS (red).

**Table 1.** Names and top active codes of the 40 topics inferred by treeLFA on the top-436 UKB dataset. Top ICD-10 codes (with an unnormalized probability of at least 0.3) in the 40 topics inferred by treeLFA on the top-436 UKB dataset are shown. Topics are named using the ICD-10 chapters which make major contributions to their top active codes (different chapters are connected with “-” in the names). Words after colons in the names of topics give further summary of the top active codes from a specific chapter. Topic 1 and 2 are not named since they are very dense, and there is no obvious pattern in their top active codes.

| Topic | Name | Top active codes |
| --- | --- | --- |
| 1 | - | - |
| 2 | - | - |
| 3 | **Musculoskeletal:joint-1** | M19,I10,M17,M25,M13,M54,K21,E78,M47,M23,K44,E66,M15,M79,J45,K29,M16,M75,K57,G56,F32,E11,M51,M65 |
| 4 | **Neoplasm-Digestive** | C78,C18,C77,I10,K56,D64,K63,C79,K59,K57,K66,C20,J90,K91,N39,K62,J18,K52,E87,N17,D12,B96,K43,K29,E86,A41 |
| 5 | **Neoplasm-Respiratory-Blood** | C79,C78,D70,C77,J18,A41,J90,C34,I10,D64,J22,K59,C50,E87,N17,N39,A09,C80,J98,K52 |
| 6 | **Digestive-Circulatory** | I10,I25,E78,K29,I20,K44,K57,K21,E11,K62,K63,I84,D12,K22,D50,J45,D64,K20,K92,J44,K31,I48 |
| 7 | **Nervous:cerebrovascular** | I10,I63,E78,G81,I67,N39,I69,G40,F32,I48,J18,K59,E87,H53,B96,G45,J22,G93 |
| 8 | **Respiratory-Mental** | J44,J18,F17,F32,J45,I10,F10,J22,F41,J43,J98,J47,J96,E87 |
| 9 | **Circulatory:heart-Metabolic-1** | I10,I25,I48,I50,E78,I51,I20,I44,I34,I21,I08,I47,I35,J90,J18,I49,I42,E11 |
| 10 | **Urinary:male-1** | I10,N32,N40,N39,E78,C67,N30,N35,N13,E11,N20,C61,K57 |
| 11 | **Metabolic:diabetes-Eye** | E11,I10,E78,E10,H26,H36,E14,I25,H25,E66 |
| 12 | **Musculoskeletal:spine** | M51,M54,G55,M47,M48,I10,M79,M43,M50 |
| 13 | **Reproductive:female-1** | D25,N83,N92,N80,N73,N94 |
| 14 | **Skin:infection** | L03,I10,L40,B95,L02 |
| 15 | **Musculoskeletal:joint-2** | M19,M20,M25,I10,M75,M16 |
| 16 | **Reproductive:female-2** | N95,N84,N85,I10,D25,N81 |
| 17 | **Eye-1** | H26,H25,I10,H35,H40,H33,H52 |
| 18 | **Circulatory:heart-Metabolic-2** | I25,I10,E78,I20,I21 |
| 19 | **Digestive:lower GI-1** | K52,K51,K62,K63,K50,K57,I84,A09 |
| 20 | **Digestive:lower GI-2** | K57,D12,K63,K62,I84,I10 |
| 21 | **Digestive:upper GI** | K44,K29,K21,K22,K20,K31,I10 |
| 22 | **Digestive:hepatobiliary** | K80,I10,K81,K82,K85 |
| 23 | **Urinary:female** | N39,N81,N32 |
| 24 | **Neoplasm:skin-Skin** | C44,D22,L57,L98,L82,I10,C43 |
| 25 | **Urinary:male-2** | N40,N32,I10,C61,K40 |
| 26 | **Metabolic** | I10,E78,I48,E11 |
| 27 | **Renal:kidney** | N20,I10,N23,N13 |
| 28 | **Respiratory:upper** | J34,J33,J32,J45 |
| 29 | **Ear:hearing** | H72,H91,H90,H65,I10 |
| 30 | **Musculoskeletal:knee** | M23,M17,I10 |
| 31 | **Neoplasm:breast** | C50,C77,D05 |
| 32 | **Digestive:lower GI-3** | I84,K62,K57 |
| 33 | **Reproductive:female-4** | N92,D25,N84 |
| 34 | **Circulatory:peripheral**<br>**vascular** | I80,M79,I83,I26 |
| 35 | **Digestive:teeth** | K02,K08,K01,K04 |
| 36 | **Reproductive:female-5** | N87,N84,N95 |
| 37 | **Eye:appendicle** | H02,H04 |
| 38 | **Eye-2** | H26,H33 |
| 39 | **Digestive:hernia** | K40 |
| 40 | **Healthy** | Empty |

We then performed topic-GWAS on the 40 treeLFA and flatLFA topics we kept. We found 278 treeLFA and 260 flatFLA genome-wide significant loci, with the majority (207) found in both sets and associated with corresponding topics (Supplementary Figure 10c-d, Supplementary Table 16). We also performed single-code GWAS on the 436 ICD-10 codes and found 1,093 associated loci, among them 198 were also associated with treeLFA topics. Lead SNPs for loci only associated with treeLFA topics (80 unique loci) had smaller effect sizes (median absolute effect size: 0.021) compared to loci supported by both topics and single codes (0.024) (Figure 6d, Supplementary Table 17), indicating that topic-GWAS enabled the discovery of variants with small effects on multiple related diseases. Unique loci were not uniformly distributed across topics; as in the top-100 dataset, many were associated with the empty topic (21). Topics 3 and 30 also have large proportions of unique loci (81.5% and 83.3%), and most of their active codes are from Chapter 13 (Diseases of the musculoskeletal system and connective tissue) (Figure 6e, Supplementary Table 18). Other topics that are associated with substantial numbers of unique loci are shown in the Supplementary Table 19.

We validated the topic-associated loci for the larger dataset using the same approach as used with the top-100 dataset. Overall, 89.2% (248/278) of topic-associated loci and 78.9% (63/80) of unique associations have records in the GWAS catalogue. The functional validation results were also similar to that on the top-100 dataset, with unique topic-associated loci and single code associated loci exhibiting similar profiles (Supplementary Figure 11). The two types of PRS for single codes were also constructed using single code and topic-GWAS results. For 130 in 436 (30%) ICD-10 codes, PRS based on topic-GWAS resulted in larger AUC on the test data. These codes were mainly from Chapter 3 (Diseases of the blood and blood-forming organs and certain disorders involving the immune mechanism, 5/12), Chapter 9 (Diseases of the circulatory system, 21/48) and Chapter 13 (Diseases of the musculoskeletal system and connective tissue, 24/52). In contrast, for most (93 of 109) ICD-10 codes with more than 5 associated loci, PRS based on single code GWAS resulted in better performance (Supplementary Table 20). To compare treeLFA and flatLFA, we also constructed PRS for single codes based on flatLFA results, and compared their AUC on the test data to that of treeLFA. For 231 in 436 codes (53%), PRS based on treeLFA showed better performance. Supplementary Figure 10e compares the density plots for the AUC of PRS for all codes given by the two methods, where treeFLA shows a minor advantage.

## Discussion

Multimorbidity is a major challenge for today’s healthcare systems, yet our understanding of it remains limited ^58^. The establishment of biobanks linked to electronic health records presents an opportunity for a more systematic study of multimorbidity, and highlights the need for reliable and powerful analytic tools to enable the identification of major multimorbidity clusters and downstream analyses of paired phenotype and -omics data.

Here we developed treeLFA, a topic model inspired by Latent Dirichlet Allocation (LDA) that admits a prior for topics constructed on existing tree-structured medical ontologies. We compared it to flatLFA and LDA on both simulated and UKB data, and found that the prior was effective at extracting relevant topics from limited input data, such as data involving rare diseases. We also found that the novel model structure better fits the binary input data, resulting in the identification of an empty (“healthy”) topic, and ensuring that topic weights for the remaining disease topics were largely uncorrelated, improving power for downstream topic-GWAS. We implemented algorithms to optimise hyperparameters of the model, and developed a computationally efficient approach to determine the number of meaningful topics to be inferred.

By applying treeLFA to HES data for 436 common diseases recorded in UKB, we identified 40 topics reflecting combinations of diseases that tend to co-occur. These topics varied in density and include a single empty topic, many sparse topics that each include a small number of active disease codes, and several dense topics. We found that the inferred topics were largely consistent with the current disease classification system (ICD-10), yet treeLFA also combines diseases distant on the tree structure into the same topic, indicating the utility of supplementing expert-led knowledge systems with data-driven methods. By comparing the topics inferred by treeLFA and LDA, and topics inferred by different treeLFA models, we found that multimorbidity clusters were consistently inferred, indicating the existence of stable multimorbidity clusters, and that topic modelling of cross-sectional diagnosis data is an informative method of finding them.

Most inferred topics likely reflect underlying aetiology, as indicated by the fact that the large majority (34/40) show genome-wide significant associations with genetic markers, while 20 topics are associated with 80 novel loci that do not reach genome wide significance in GWAS for single ICD-10 codes, and show evidence of functionality using multiple methods. The active ICD-10 codes in the topics with the most novel associations are mainly from Chapter 13 (Diseases of the musculoskeletal system and connective tissue), with substantial contributions also from Chapters 4 (Endocrine, nutritional and metabolic diseases), 9 (Diseases of the circulatory system) and 14 (Diseases of genito-urinary system), suggesting that diseases in these chapters share genetic risk factors.

With topic-GWAS results, we explored constructing PRS for a single code as the sum of PRS for all topics weighted by the probabilities of the code in topics. We found that for certain codes, especially codes with very few GWAS hits from Chapter 13, this new type of PRS outperforms the standard PRS based on single code GWAS results. This improvement in prediction might result from better estimation of effect sizes of variants by topic-GWAS. This is because although treeLFA factorises the input matrix in a linear way, it achieves a dimension reduction by mapping from the disease space to the topic space. This results in fewer traits and therefore more ‘cases’ for each one, which is especially beneficial for the study of the highly polygenic traits (topic weights), where there are a large number of SNPs with small effects.

In contrast to topic-GWAS, single-code GWAS on the 463 common diseases resulted in 1,093 significant associations, of which the vast majority (895) were not associated with any topic. Taken together, these genetic analyses indicate that the majority of genetic associations are driven through links to individual diseases. Meanwhile, most multimorbidity clusters also have genetic bases, which are mainly composed of pleiotropic genetic variants affecting risks of multiple active diseases in the cluster. Besides, there are also a substantial number of associations for topics that are difficult to identify by single code GWAS due to the lack of power. From a biological perspective, they might reflect the complex connections between upstream pathways that are distant to individual diseases, or variants with direct yet small effects across multiple diseases. Overall, these observations are a helpful starting point for our pursuit of a deeper understanding of the mechanisms underlying multimorbidity clusters.

We consistently identified an “empty” topic, which reflects individuals’ overall disease burden, since their weights for the “empty” topic are negatively correlated with their total number of diagnosed diseases. Perhaps surprisingly we found that this topic showed many genetic associations, many of which (21/35) had not been identified before. An enrichment analysis for topic-associated genes (Methods) indicated several lifestyle-associated factors, such as gym attendance and religious observance, suggesting that this topic may be related to individuals’ health behaviour, which in part is genetically determined. In contrast, for most sparse disease topics, the enriched gene sets are usually directly related to the active codes in the topic (Supplementary Table 21).

There have been many studies aiming at identifying multimorbidity patterns using various methods ^12, 13, 16, 17, 59^. Most of these studies focus on dozens of diseases that in addition varied from study to study, making comparisons difficult. We compared the topics identified by treeLFA with multimorbidity networks (which are interpretable at the level of genetic loci) found by a recent study using the HES data for 439 common diseases in UKB (433 of these are included in the top-436 UKB dataset in our study) ^11^. For most of the disease networks, there are specific corresponding disease topics identified by treeLFA (Supplementary Table 22). However, the overlap of active codes in treeLFA inferred topics and the disease networks is limited, suggesting significant differences in the details of the inference results. This discrepancy could be caused by the fundamental differences between the two methods, since treeLFA analyses all diseases simultaneously, while multimorbidity networks were constructed based on pairs of diseases that tend to co-occur. One limitation of topic models is that they cannot determine the relationships between active diseases in the same topic. In contrast, an advantage of topic models is that they make direct use of individual level data for the genetic analyses, and allows for making predictions on the test data, which provides an objective way to compare different methods.

Our work represents real progress in the understanding of multimorbidity, yet also reveals important and unsolved challenges. For instance, while we showed that taken together the novel genetic associations likely represent true biology, we have not performed individual replication of the findings in this study in independent data sets and this may be challenging unless the data sources and methods of data collection are comparable to UK Biobank. The problem of inferring disease topics that are stable, tractable and biologically meaningful across geographies and healthcare systems represents a major challenge for future research.

## Supporting information

Analytic note

## Data Availability

This research has been conducted using the UK Biobank Resource; application number 12788. 

## Acknowledgements

This work uses data provided by patients and collected by the NHS as part of their care and Support. Funded by the Chinese Academy of Medical Sciences (CAMS) China Oxford Institute (COI) and Studentship from the China Scholarship Council (CSC) (YZ).

Computation used the Oxford Biomedical Research Computing (BMRC) facility, a joint development between the Wellcome Centre for Human Genetics and the Big Data Institute supported by Health Data Research UK and the NIHR Oxford Biomedical Research Centre. We thank Chris Holmes for the discussion.

## Author Contributions

Y.Z.: conceptualization, data curation, formal analysis, methodology, software, validation, visualisation, writing-original draft; X.J.: conceptualization, data curation, methodology, software, writing: review & editing; A.J.M: conceptualization, project administration, supervision, writing: review & editing; G.L: conceptualization, investigation, methodology, project administration, resources, supervision, writing-original draft, writing: review & editing; GM: conceptualization, funding acquisition, methodology, project administration, resources, supervision, writing: review & editing;

## Methods

Full technical details for treeLFA are given in the analytical note in the supplemental data.

### Validation of treeLFA with simulated data

The simulation study served two purposes. Firstly, we aimed to verify that treeLFA can accurately infer latent topics from the diagnosis data encoded as binary variables. Secondly, we aimed to compare the performance of the three topic models (treeLFA, flatLFA and LDA) discussed in the next section, such that the influence of model structure and treeLFA’s informative prior for topics on the inference can be assessed.

### Overview of the three related topic models

#### 1. treeLFA

treeLFA (“latent factor allocation with a tree structured prior for topics”) is a topic model designed for binary diagnosis data in biobanks based on the Bayesian mean-parameterized binary non-negative matrix factorization ^41^. It models the presence and absence of S disease codes for D individuals with D×S Bernoulli distributions, and the matrix of Bernoulli probability for all disease variables is factored into the topic matrix (ϕ) and the topic weight matrix (θ). The loading of disease code s in topic t (ϕ_ts_) is its corresponding Bernoulli probability in the topic, and the Bernoulli probability of the disease variable for individual d and disease code s is a mixture of the Bernoulli probability of code s in all topics, with the mixing coefficients specified by the topic weights for individual d. treeLFA also incorporates an informative prior for topics constructed by running a Markov process on a tree structure of individual words, which assumes that disease codes on the same subtree are likely to have similar Bernoulli probability in the same topics (see the details in the analytic note).

#### 2. flatLFA

flatLFA has the same model space as treeLFA, with the only difference being that flatLFA uses a non-informative prior for topics, which is constructed on a tree structure where all nodes (representing all disease codes in a topic) are placed directly under the common root node. By comparing the performance treeLFA and flatLFA, the contribution of treeLFA’s informative prior for topics to the inference can be assessed.

#### 3. LDA

Latent Dirichlet Allocation (LDA ^60^)’s model configuration is different from treeLFA. LDA only models the disease codes that are diagnosed for individuals with categorical distributions. For LDA, each topic is a categorical distribution (or a Multinomial distribution if the input data is viewed as a count matrix) across the S disease codes, and a Dirichlet prior distribution is used to generate these topics. By contrast, for treeLFA each topic is a sequence of Bernoulli probability for the S disease codes, and S Beta distributions are used to generate the topics.

#### Description of simulated data

We simulate multiple data sets using different topics and hyperparameters to assess the performance of the three topic models (treeLFA, flatLFA and LDA) in different situations. We simulate the input data sets in two steps. Firstly, we build a tree structure for 20 disease codes (Figure 2B). The tree structure has three layers. The first layer is the root node; the second layer contains five nodes, and each of them has three children nodes in the third layer. Secondly, we generate topics of disease codes using the tree structure above. A Markov process on the tree structure is used to do this (see the analytic note), and the rationale for choosing the parameter of the Markov process is explained below.

The Markov process chooses active disease codes (disease codes having large probability in a topic) for each topic by generating binary indicator variables for disease codes (with 1 represents active codes, and 0 represent inactive codes in a topic) (see the analytic note). The Markov process has two transition probabilities, ρ_01_ and ρ_11_, which control the sparsity of topics (ρ_01_ = P(I = 1|I^parent^ = 0), ρ_11_ = P(I = 1|I^parent^ = 1)). Small values for both ρ_01_ and ρ_11_ give rise to sparse topics (because most codes in a topic will be inactive), while large values for both generate dense topics. ρ_11_ also controls the clustering of active codes in topics. With a large ρ_11_, most children codes of an active parent code will be active. As a result, active codes in a topic will gather on the same branch of the tree. By contrast, if ρ_11_ is small, active codes will spread across the entire tree. For our simulation, we use topics resembling those generated with small ρ_01_ and large ρ_11_. This reflects our belief that in the real world most topics of disease codes should be sparse (thus we chose small ρ_01_), and that active disease codes in the same topic tend to come from the same subtree (thus we chose large ρ_11_).

For our simulation, we construct two sets of topics manually. The first set of topics are likely to be generated using a Markov process with small ρ_01_ and large ρ_11_ (hyperparameter setting used for inference on the simulated dataset), while the second set of topics are unlikely to be generated by this Markov process. The first set of topics are used to test if the tree structure of codes improves inference accuracy, and the second set of topics are to test the robustness of treeLFA’s inference when the tree structure of codes is wrongly specified. We manually specified the topics to ensure that they are completely distinct from each other and have strong patterns with respect to the clustering of active codes.

Figure 2B shows the first set of topics. For the first three topics, all codes on one branch of the tree are active, and the remaining codes are inactive. In the last topic, all codes from two branches of the tree are active. These topics are likely to be generated using a Markov process with small ρ_01_ and large ρ_11_, since a parent code and all its children codes are always in the same state (either active or inactive). In the second simulation setting, active diseases in topics are not generated according to their adjacency on the tree (Supplementary Figure 1B). We construct these topics by switching a fraction of active codes between topics in the first simulation setting. As a result, active parent codes always have inactive children codes, and inactive parent codes always have active children codes.

We simulate disease data using the topics described above and the generative process of treeLFA. For each dataset, we split the data into training and testing data of the same size. To evaluate the topic models in different situations in each topic setting, we use four combinations of two hyperparameters to simulate data: α (the concentration parameter of the Dirichlet prior for topic weights θ) and D (the number of individuals in the training dataset). A large value for α means that most individuals will have large topic weights spread across topics. By contrast, a small value for α will make topic weights for each individual more concentrated on a single topic. A large α makes the inference difficult, since most individuals are a mixture of multiple topics. Therefore, data sets simulated using large α require larger D for accurate inference. For each hyperparameter and topic setting, we simulate 20 datasets. Supplementary Table 1 summarises the hyperparameter and topic settings.

#### Implementation of the inference procedure

LDA is implemented using the R package “topicmodels”, and collapsed Gibbs sampling is used to do the inference. treeLFA and flatLFA are implemented from scratch by us using the R package “RcppParallel” and “Rcpp”.

For the hyperparameters of treeLFA and flatLFA, we provide the true value of α to train the three models. Beta priors for the probability of active and inactive disease codes in topics (ϕ) are Beta(2,4) and Beta(0.3,80). Beta priors for the transition probability (ρ_01_ and ρ_11_) of the Markov process are Beta(4.8,20) and Beta(20,4.8) (treeLFA). For flatLFA only ρ_01_ will be used on the tree, and its prior is Beta(7,20), resulting in approximately the same expected number of active codes in topics as treeLFA. For LDA we try a few different values (0.01,0.1,1) for η, the concentration parameter of the Dirichlet prior for topics. We find that with η = 0.01 LDA has the best performance evaluated by the inference accuracy.

For the initialization of hidden variables for treeLFA and flatLFA, we initialised all indicator variables (I) as 0, and then simulate all probability variables (ϕ) using the Beta prior for inactive disease codes. Topic assignment variables (Z) are randomly sampled for all individuals.

For each simulation scenario, ten Gibbs chains were sampled, and 20 posterior samples of hidden variables were collected from each Gibbs chain with an interval of 100 iterations after 15,000 burn-in iterations.

#### Evaluation and comparison of topic models on simulated datasets

Two metrics are used to evaluate the models in simulations. The first metric is the inference accuracy, measured with the averaged per disease difference between true and inferred topic loadings: 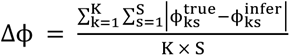, where ϕ_ks_ is the Bernoulli probability of disease code s in topic k. To reorder the inferred topics as the true topics, we match each inferred topic to the true topic that has the highest cosine similarity, in a greedy procedure (i.e. once a true topic is matched, it is removed from the matching of the next inferred topic). The pairwise t-test is used to test for statistical difference between two different models. The second metric is the predictive likelihood on the test data (see the analytic note for more details). For each posterior sample of topics, 200 Monte-carlo samples of topic weight θ are used to approximate the predictive likelihood. A sensitivity analysis is done to ensure the number of samples for θ is enough to have a stable estimate of the predictive likelihood.

### Inference on the top-100 UK Biobank dataset

#### The input data for treeLFA

The input to treeLFA includes the diagnosis data for individuals in the UK Biobank, and the tree structure for disease codes. The diagnosis dataset is constructed from the Hospital Episode Statistics (HES) data in the UK Biobank, which is coded using the five-layered hierarchical ICD-10 billing system. The first layer of the ICD-10 tree structure is the root node; the second layer is composed of chapters of diseases coded using capital English letters; the third layer contains blocks of disease categories; the fourth layer contains single disease categories; and lastly, the bottom layer contains sub-categories of diseases, which can be, for instance, the same disease occurring at different sites of human body, or subtypes of the same disease. In UK Biobank, most of the diagnosed diseases are encoded using codes on the bottom layer (fifth layer) of the tree. We use the fourth layer of encoding as diagnoses, where we replace all diagnoses with their parental code in the fourth layer.

The top 100 most frequent ICD-10 codes in UK Biobank from the first 13 chapters of the ICD- 10 coding system are chosen to construct the dataset. This selection of chapters provides a balance between breadth of phenotype and depth within any one chapter so that the potential benefits of treeLFA can be explored. The diagnosis data is a binary matrix, with each row represents an individual, and each column a disease code. Zeros and ones in the matrix are used to represent the absence and presence of diagnosed ICD-10 codes for individuals. If an individual is diagnosed with the same disease code several times, we keep only one record to avoid bias of repeated diagnoses. The full dataset is randomly split into a training dataset and a testing dataset, containing the diagnosis record for 80% and 20% individuals.

The tree structure of disease codes is encoded in a table with 2 columns: the first column contains all the ICD-10 codes on the tree, and the second column records the parent codes of the corresponding codes in the first column (Supplementary Table 3).

#### Implementation of treeLFA

##### Training strategy for treeLFA

The Gibbs-EM algorithm is firstly used to optimise α in two stages. In the first stage we run 2,000 iterations of the Gibbs-EM algorithm. In the E-step of each iteration, we run the Gibbs sampler for treeLFA for 20 iterations and collect one posterior sample of Z (19 burn-in Gibbs sampling iterations before the collection of the posterior sample). In the M-step, α is optimised using this single posterior sample of Z collected in the E-step. In the second stage we continue to run the Gibbs-EM algorithm for 200 iterations. In the E-step of each iteration, we run the Gibbs sampler for treeLFA for 200 iterations and collect ten posterior samples of Z in total (19 burn-in Gibbs sampling iterations before the collection of each posterior sample). The reason to have two stages of training is to balance the computational speed with the inference accuracy. In the first stage, we optimise α more frequently and quickly get close to its optimal value. In the second stage, α is more accurately optimised based on multiple posterior samples of Z.

After optimising α, we use collapsed Gibbs sampler to simulate posterior distributions of all hidden variables (Z, I, ϕ, ρ), with α fixed at and hidden variables initialised at the values provided by the last iteration of the Gibbs-EM algorithm. 5,000 burn-in iterations are run before the collection of posterior samples of hidden variables. For each topic model, ten Gibbs chains are constructed, and 50 posterior samples are collected with an interval of 100 iterations from each chain.

##### Choices of hyperparameters and initialization of hidden variables

To shorten the training with the Gibbs-EM algorithm, we initialise α as (1,0.1,…,0.1). The first entry in α is much larger than the others, and it corresponds to the empty topic that will always be inferred from real-world diagnosis data. We also initialise α in other ways, such as using (1,…1), and we find that model converge to the same results regardless of the ways of initialization of α, and the optimised α is usually more close to (1,0.1,…,0.1) than other choices. For topic assignment variable Z, we assign the empty topic (topic 1, corresponds to the first entry in α) to all disease variables for individuals without any diagnosed disease codes. For individuals with at least one diagnosed disease code, all topics are randomly assigned to all disease variables. For topics, all indicator variable I are initialised as 0, and probability variable ϕ are randomly sampled from Beta(1,5,000,000). Beta(0.3,80) and Beta(2,4) are used as the prior for ϕ of inactive and active codes. Beta(3,20) and Beta(3,3) are used as the prior for transition probability ρ_01_ and ρ_11_ of the Markov process on the tree. The hyperparameters for flatLFA are set in the same way as treeLFA. For LDA, the concentration parameters of the Dirichlet priors for topic weights (α) and topics (η) are both initialised as a vector of 0.1. α is not optimised since we find that this has negligible influence on the inference result (inferred topics) and downstream analyses (topic-GWAS).

#### Post-processing of inference result

##### Approximation of topic weights

The topic weight variable θ is integrated out during the collapsed Gibbs sampling, therefore their posterior samples need to be approximated using posterior samples of Z and α. θ can be computed as in Griffiths and Steyvers ^53^: 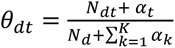, where N_dt_ is the total number of disease variables assigned with topic t for individual d, and N_d_ is the total number of disease variables.

##### Combining inference results from different Gibbs chains

To combine the inference results given by different Gibbs chains, the “identifiability” issue needs to be addressed, since the order of topics in different posterior samples from different chains may not be the same.

We combine all posterior samples of topics from all chains together, and cluster topics before taking the average within each cluster. To cluster topics from all samples, we firstly construct a shared nearest neighbour (SNN) graph using the R package “scran” ^61^. With the SNN graph, we use the “Louvain” algorithm ^62^, a community detection algorithm implemented in the R package “igraph”, to assign topics into clusters. After clustering, similar topics coming from different chains or posterior samples will be put into the same cluster. In addition to topics (ϕ), we also assign posterior samples of other hidden variables (I, ρ and α) to the corresponding clusters according to the clustering result for topics.

The Louvain algorithm doesn’t allow us to directly specify the total number of clusters (communities) to be found. Instead, the number of clusters is decided by the hyperparameter k (the number of nearest neighbours to consider) for the construction of the SNN graph. A large k will result in a small number of clusters, while a small k gives rise to a large number of clusters, though some clusters might be alike. Empirically, we choose 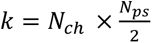, where N_ch_ is the number of Gibbs chains for the same treeLFA model, and N_ps_ is the total number of posterior samples taken from each chain. This choice is to balance the total number of clusters found by the algorithm and the uniqueness of different clusters.

##### Post-processing inference results from models with a large number of topics

For models set with a very large number of topics which far exceed the actual number needed to explain the data (for instance, the treeLFA models set with 50 or 100 topics), multiple near-empty topics (topics with few active codes having very small probability) will be inferred. Although the small differences between these near-empty topics are not meaningful, they are usually assigned to different clusters by the Louvain algorithm. To collapse these near-empty topics into a single empty topic, we further apply hierarchical clustering on topics averaged from different clusters given by the Louvain algorithm. During the hierarchical clustering, similar topics are kept being combined until all the remaining topics are significantly different from each other. By visualising the inferred topics using heatmap, one can roughly decide the number of distinct meaningful topics (topics that are not empty or near-empty) to keep, and then set the number of clusters (topics) to keep for the hierarchical clustering.

### Genetic analyses

#### topic-GWAS and single code GWAS

To find genetic variants influencing individuals’ risks for topics of diseases, we perform GWAS using inferred topic weights as continuous traits (topic-GWAS). Since topic weights are real numbers in the range of 0 to 1, the basic assumptions of linear regression do not hold. We apply a logit transformation on topic weights to address this issue before fitting the standard linear model for GWAS. We validate that using logit transformation gives better results than using rank based inverse normal transformation and using no transformation on topic weights. The validation is done by comparing the number of significant loci found by different methods, and the predictive performance of PRS for single codes based on topic-GWAS results (see section below). For topic-GWAS, we only include common SNPs (SNPs with a minor allele frequency (MAF) larger than 0.01 in the UK Biobank) and individuals who self-report having British ancestry in the training dataset (343,006 individuals in total). Sex, age and the first ten principal components (PCs) of genomic variation are controlled for.

For comparison, we also perform GWAS (logistic regression) using the presence and absence of single ICD-10 codes as binary traits (single code GWAS). The inclusion criterion for individuals, SNPs and covariates are the same as topic-GWAS. In addition to ICD-10 codes, we also use terminal Phecodes mapped from the top 100 ICD-10 codes as traits for single code GWAS. Phecodes are defined by systematically grouping terminal ICD-10 codes into more applicable medical terms based on the judgements of clinicians and researchers ^63^, which reduces the granularity of terminal ICD-10 codes. Similar to ICD-10 codes, there is also a hierarchical coding system for Phecodes. To map the ICD-10 codes used in the top- 100 UKB dataset to phecodes, we firstly extract all terminal ICD-10 codes (on the fifth layer of the ICD-10 tree) that are children codes of the 100 level-4 ICD-10 codes, and then retrieve their corresponding Phecodes according to the Phecode map (https://phewascatalog.org/phecodes) In total, there are 296 terminal Phecodes mapped from the 100 ICD-10 codes.

##### Inflation in P-values given by topic-GWAS

Inflation in P-values are observed for the topic-GWAS results given by all three topic models. The inflation can either be resulted from true polygenicity of the traits (topic weights), or stratification in the population. To differentiate these two possibilities, we carry out the LD score regression (LDSC) ^64, 65^ using the summary statistics of topic-GWAS for all topics. Pre- computed LD scores (based on 1000 Genomes European data) are downloaded and used in the analyses as recommended ^65^. The genomic control inflation factor λ_GC_ and the intercept of LDSC are output by the algorithm, and compared with each other. A large λ_GC_ and small intercept for the same trait suggest true polygenicity causing the inflation in P-values, while large values for both λ_GC_ and intercept suggest stratification in the population.

##### Processing GWAS results

To define genomic loci from significant SNPs (P<5×10^−8^) found by GWAS, we use the clumping function implemented in PLINK-1.9. r^2^>0.1 is used as the threshold for clumping SNPs in linkage disequilibrium (LD). We define a loci to be an association for both topic-GWAS and single code GWAS as follows: the significant lead SNP found by one GWAS method can be clumped with a significant lead SNP found by the other method.

##### GWAS on internal disease codes on the tree

In addition to grouping disease codes via topic modelling, we also group disease codes completely following the medical ontologies (ICD-10 and Phecode systems). In other words, we use internal codes (such as blocks of categories of diseases and chapters of disease, corresponding to the nodes in the third and second layers of the ICD-10 coding system) of the two disease classification systems as binary traits for single code GWAS. For instance, if both disease codes A and B are under a common parent code C on the tree, then C will be used as the trait for GWAS, and individuals who are diagnosed with either A or B will be used as cases for the single code GWAS for code C. For the 100 ICD-10 codes there are 68 internal codes, and for the 296 Phecodes there are 136 internal codes.

##### Comparison of topic-GWAS results for the three topic models

In addition to topics inferred by treeLFA, topic-GWAS for flatLFA and LDA inferred topics are also performed. For LDA, only individuals with at least one diagnosed disease code are used as input for inference. For topic-GWAS, there are two options to deal with the individuals without any diagnosis. We can either exclude them or include them and give them small random weights for all disease topics. We experiment with both methods, and find that excluding the completely healthy individuals results in a larger power for topic-GWAS.

#### Validation of topic-associated loci

##### Validation using the GWAS Catalogue

We check the GWAS Catalogue ^56^ to see if topic-associated loci were also found by previous GWAS as significant. We download the full GWAS Catalogue ^66^, and clump all SNPs in it to topic-associated lead SNPs (r^2^>0.5 as threshold). If a topic-associated lead SNP found by us can be clumped, it means that a SNP in LD with it was found by a previous GWAS as significant.

##### Validation using functional genomic resources

Integrated analysis of GWAS results and functional genomic datasets has gained popularity in recent years ^56, 67^. Checking the enrichment of genomic annotations among topic- associated loci (lead SNPs) is another angle of validation. We obtain various genomic annotations for topic-associated loci using the software “FUMA” ^68, 69^. Since most topics only have a small number of associated loci, we combine all loci (lead SNPs) that are associated with at least one topic and perform analyses on them as a whole. Meanwhile, we also perform the same analyses on all single code associated lead SNPs and 10,000 random SNPs sampled from all SNPs used in the GWAS (the distribution of their MAF are matched to all topic-associated SNPs) for comparison. The assumption made here is that if topic-GWAS find true associations, then the significant SNPs should have an enrichment profile that is similar to single code associated SNPs (positive control) and different from randomly selected SNPs (negative control).

Three types of functional annotations are used for the validation of topic-associated lead SNPs. Firstly, the three groups of loci (lead SNPs) are annotated using the 15-core chromatin states predicted by the chromHMM algorithm ^57^. Since the predicted chromatin states in the 127 available types of tissues are different, for each genomic locus we use the smallest chromatin state across all tissues. Secondly, we calculate the proportions of lead SNPs in the three groups that are eQTL (expression quantitative trait loci) in different tissues using the eQTL mapping function implemented in FUMA, based on the GTEv8 dataset ^70, 71^. Thirdly, we calculate the proportions of lead SNPs having chromatin interactions with other genomic regions in different tissues, based on the HiC data from the GSE87112 dataset ^70^. The default setting of FUMA for parameters is used in all the analyses. The two proportion z-test is used to test for significant differences between the proportions of two groups.

##### Genetic risk prediction based on topic-GWAS results

###### PRS for topics

Another way to validate topic-GWAS results is to use them for prediction tasks on the test data. Because individual variants’ effects on traits of interest are usually small, polygenic risk scores (PRS) are constructed to aggregate the effects of tens of thousands of variants. With topic-GWAS carried out on the training data, PRS for topic weights (traits of topic-GWAS) are constructed using the software “PRSice-2” ^72^, which uses a “C+T” (clumping and thresholding) method. No threshold for P-values is manually set for the inclusion of SNPs.

We use the test data to evaluate PRS for topics constructed on the training data. Topic weights for individuals in the testing dataset are inferred by running the Gibbs sampler for treeLFA on them, with ϕ and α fixed at values learnt from the training data (averaged from all posterior samples from all Gibbs chains). Ten Gibbs chains are simulated to infer topic weights for individuals in the testing dataset, and 50 posterior samples are collected from each chain, and their average is used in the subsequent analyses. With inferred topic weights, linear models are fit to evaluate the associations of PRS for topics and the corresponding topic weights, using the logit transformed topic weights as response variables, and PRS for topics as independent variables. The heritabilities of topic weights are estimated using LDSC as a reference.

###### PRS for single codes based on topic-GWAS results

To evaluate topic-GWAS results using single code GWAS results as reference, and to compare the topic-GWAS results for different topics models (such as treeLFA and LDA) under a common criterion, we construct two types of PRS for single ICD-10 codes using single code and topic-GWAS results, respectively. PRS based on single code GWAS are constructed in the standard way. As for PRS based on topic-GWAS, for code s we extract its probabilities in all topics (ϕ_ts_), and calculate an individual’s PRS for it as: PRS = 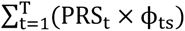, where PRS_t_ is the individual’s PRS for topic t (constructed using the topic-GWAS result for topic t). The area under the receiver-operator curve (AUC) is used to evaluate the predictive performance of PRS on the test data.

### Analyses on the larger UKB dataset

#### The input data

The larger UKB dataset (top-436 dataset) is constructed in the same way as the top-100 UKB dataset, and contains the diagnostic records of the top 436 most frequent ICD-10 codes from the first 14 chapters of the ICD-10 coding system for all individuals in UKB. These codes are all the ones in UK Biobank with a prevalence of at least 0.001 at the date of selection (continued data collection means that prevalence will tend to increase over time), corresponding to approximately 500 cases. The prevalence threshold of 0.001 is chosen both for computational reasons (this is roughly the limit of what can be performed using available computing resources) and because there must be sufficient occurrences of diseases from which to discover multi-morbidity clusters. As with the top-100 dataset, we partition the full top-436 dataset into training (80%) and testing (20%) datasets. The top-436 and the top-100 datasets use different partitions for the training and testing datasets.

#### Inference on the top-436 UKB dataset

##### Training strategy for the top-436 dataset

The top-436 dataset is more than three times larger than the top-100 dataset, increasing the computational requirements for training topic models. On the top-100 dataset, treeLFA models with different numbers of topics are trained and compared. We find that when we set an excess number of topics for the model, both inferred topics and topic-GWAS results are stable across different models (Figure 5). Therefore, on the top-436 dataset, instead of training many models with different numbers of topics, we train treeLFA and flatLFA models with 100 topics, and cluster and collapse the inferred topics to combine all near-empty topics into a single one.

For the optimization of α, the two-stage training strategy with the GibbsEM algorithm is used again. 1,500 iterations are run in the first stage (with a single posterior sample of Z collected in the E-step), and 350 iterations are run in the second stage (with 10 posterior samples of Z collected in the E-step). 50 posterior samples of hidden variables are collected during the last 50 iterations for Gibbs-EM (with an interval of 200 iterations for the Gibbs sampling). For both treeLFA and flatLFA, three Gibbs chains are simulated.

Choice of hyperparameters and initialization of hidden variables α is initialised as (1,0.1,…,0.1). Beta(0.1,3000) and Beta(1.2,3) are used as the prior for ϕ of inactive and active codes to account for diseases with small prevalence. The rest hidden variables and hyperparameters are set in the same way as for the top-100 dataset.

##### Processing inference result

We find that different Gibbs chains for treeLFA and flatLFA give slightly different inference results on the top-436 UKB dataset, while different posterior samples from the same chain have a very high level of consistency. Considering the variability among the inference results given by different chains, instead of clustering posterior samples of topics from all chains altogether, we cluster posterior samples from different chains separately. With the averaged ϕ and α for different chains, we calculate their predictive likelihood on the test data, and for both treeLFA and flatLFA we retain the chain which has the largest predictive likelihood, and use its inference result as the input for downstream analyses. For each topic inferred by the chain with the largest predictive likelihood, we check the inference results of the other chains, and annotate the topic with the number of chains that infer them to give a reference of its reliability.

#### Genetic analyses

##### topic-GWAS for the larger UKB dataset

Since the top-436 dataset is much larger than the top100 dataset, to increase the inference accuracy for topic weights, after the training with Gibbs-EM algorithm we use Gibbs sampling to re-estimate individuals’ topic weights, which is observed to increase the power of topic- GWAS. ϕ and α are fixed at values averaged from all posterior samples from the chain with the largest predictive likelihood. As a result, there is no longer an identifiability issue, so the results given by different chains (for the re-estimation of topic weights) can be combined directly. For both treeLFA and flatLFA, ten Gibbs chains are used to re-estimate topic weights, and 50 posterior samples are collected from each chain. Topic weights averaged from these chains are used as the input for topic-GWAS.

##### Gene-set enrichment analysis for topic-associated SNPs

The software FUMA can find genes that are close to the significant SNPs found by GWAS on the genome (the physical mapping function of FUMA). With the mapped genes, further analyses can be performed. Gene-set enrichment analysis (GSEA) tests for the enrichment of different gene sets among a group of genes. We choose genes that are associated with different traits in the GWAS catalogue as the reference gene sets to carry out GSEA for genes mapped from topic associated SNPs. By doing this, we can summarise the major associations of topic-associated SNPs found by previous GWAS. The default setting for FUMA is used in all the analyses in this section.

## Resource availability

### Lead Contact

Further information and requests for resources should be directed to and will be fulfilled by the lead contact, Gil McVean.

### Materials availability

The key inference results (inferred topics) of the topic models and the topic-GWAS results are included in the supplementary material of the paper. The remaining results will be made available via the UK BioBank data return and linked to UK Biobank application number: 12788.

### Data and Code Availability

This research has been conducted using the UK Biobank Resource: application number 12788. The genotype data used for GWAS in this study comes from data field “22418” in UK Biobank. The diagnosis data used for topic modelling comes from data fields “41202” and “41204” in UK Biobank.

The code for the treeLFA algorithm and a demo for using it on example data is available at: https://github.com/zhangyd10/treeLFA-demo or https://doi.org/10.5281/zenodo.7420615.

## Supplementary Information

### Description of Supplementary Data

Supplementary information includes 11 figures, 39 tables, and the analytical note.

### Supplementary Figures

**Supplementary Figure 1.**
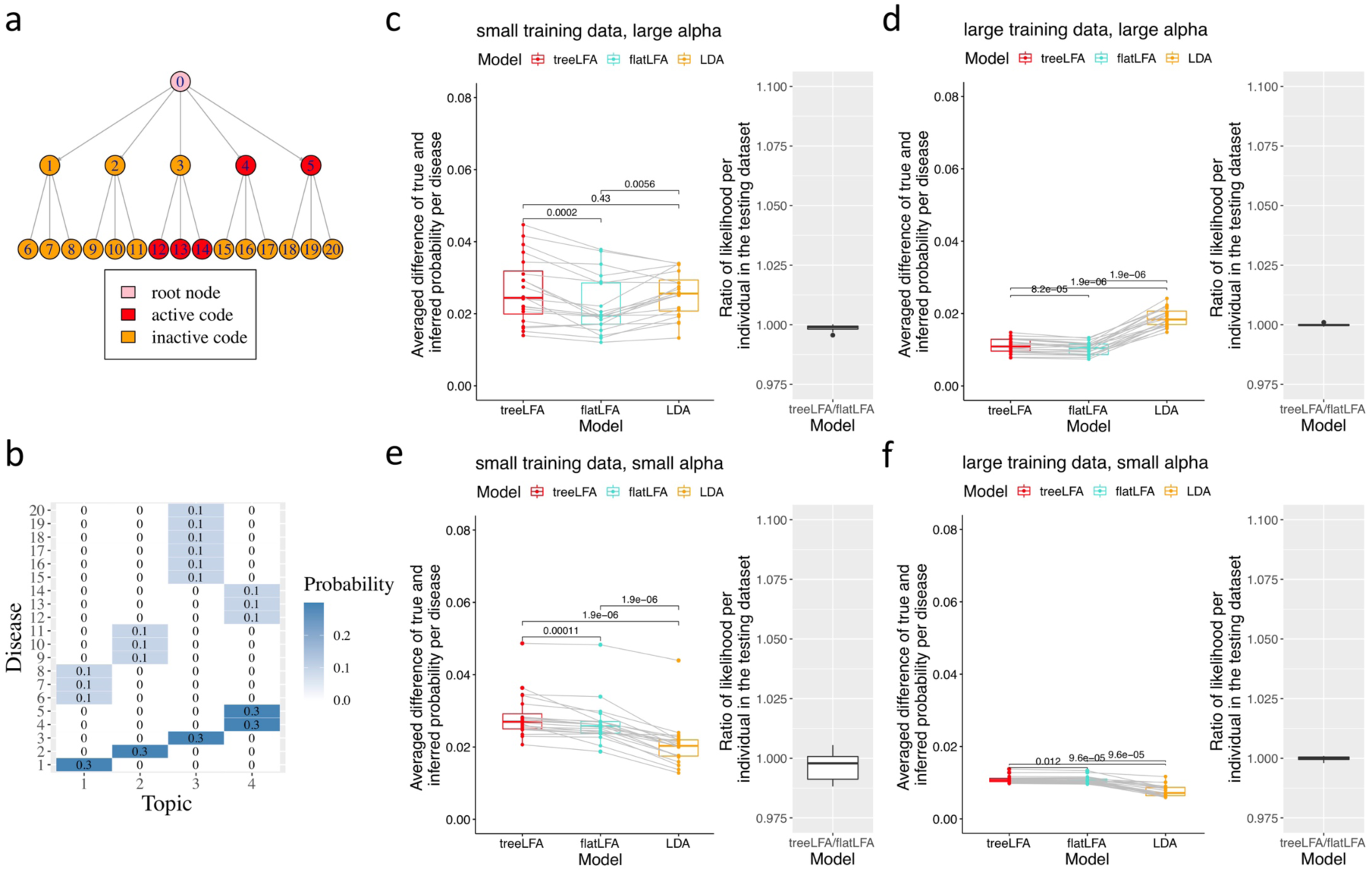
Comparison of three related topic models (treeLFA, flatLFA and LDA) on simulated datasets. a, The tree structure of 20 diseases. Red nodes correspond to the active codes in Topic 4 in panel b. b, The four topics used for simulation. Active codes in these topics are unlikely to be generated by a Markov process with small probability of transforming from inactive to active and large probability of staying active while going from the parent node to its children nodes, since an active parent code always has inactive children codes, while active children codes always have inactive parent code. c-f Comparison of three topic models on simulated datasets. The Parameter setting and metrics are the same as those in Figure 2. c, Results on datasets simulated using D=2500 and α=1. d, Results on datasets simulated using D=5000 and α=1. e, Results on datasets simulated using D=300 and α=0.1. f, Results on datasets simulated using D=1000 and α=0.1. The numeric results are in Supplementary Table 23.

**Supplementary Figure 2.**
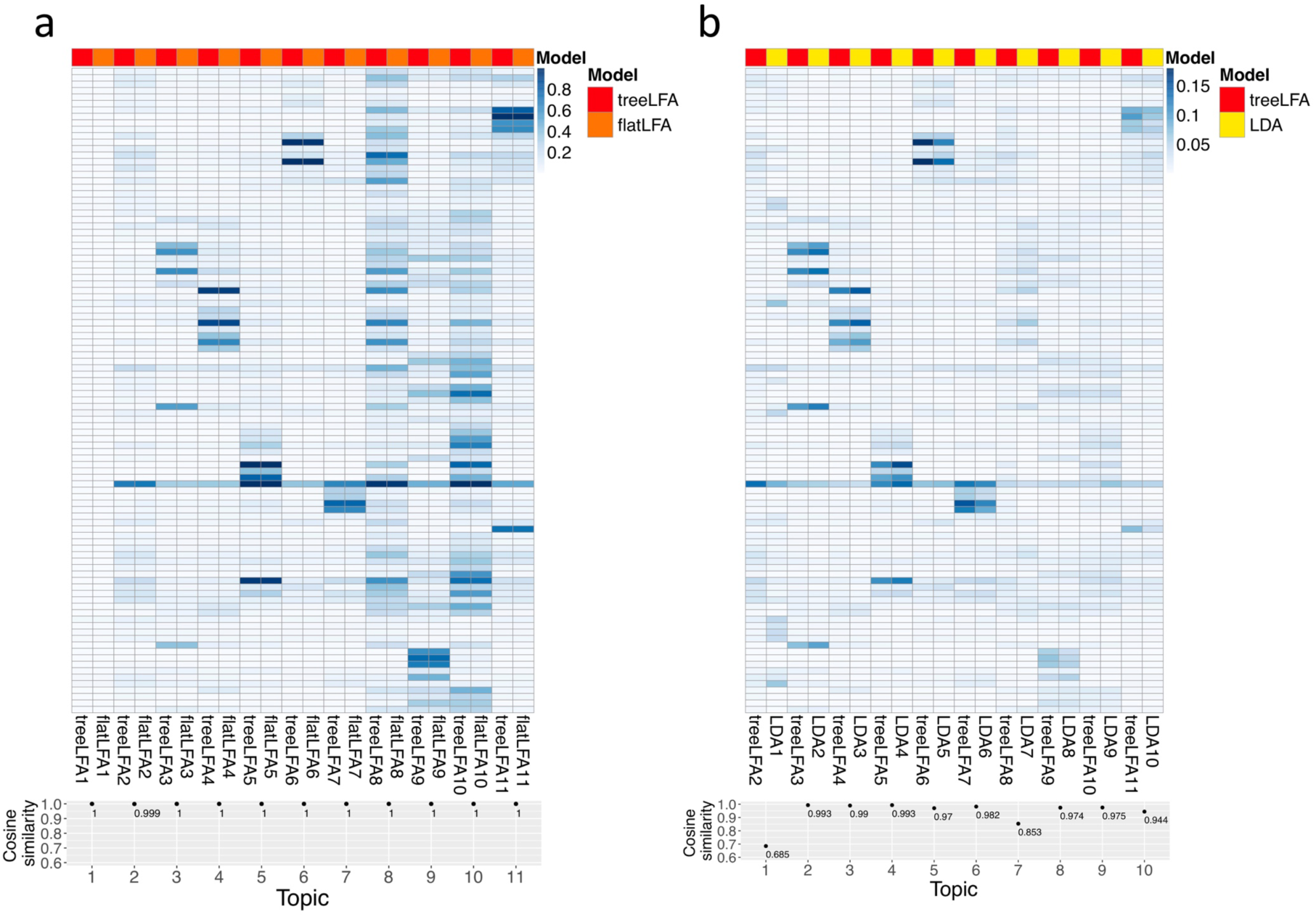
Comparison of topics inferred by three topic models on the top-100 UKB dataset. a, Comparison of the 11 topics inferred by treeLFA and flatLFA. The same topics inferred by the two models are placed next to each other. Cosine similarity was used to measure the similarity of topics inferred by the two models (point plot below the heatmap). The numeric results are in Supplementary Table 4,24. b, Comparison of the 10 topics inferred by LDA and the 10 non-empty topics inferred by treeLFA. Topics inferred by treeLFA are normalised such that probabilities of the 100 ICD-10 codes add up to 1 in any topic. The same topics inferred by the two models are placed next to each other. Cosine similarity was used to measure the similarity of topics inferred by the two models (point plot below the heatmap). The numeric results are in Supplementary Table 4,25.

**Supplementary Figure 3.**
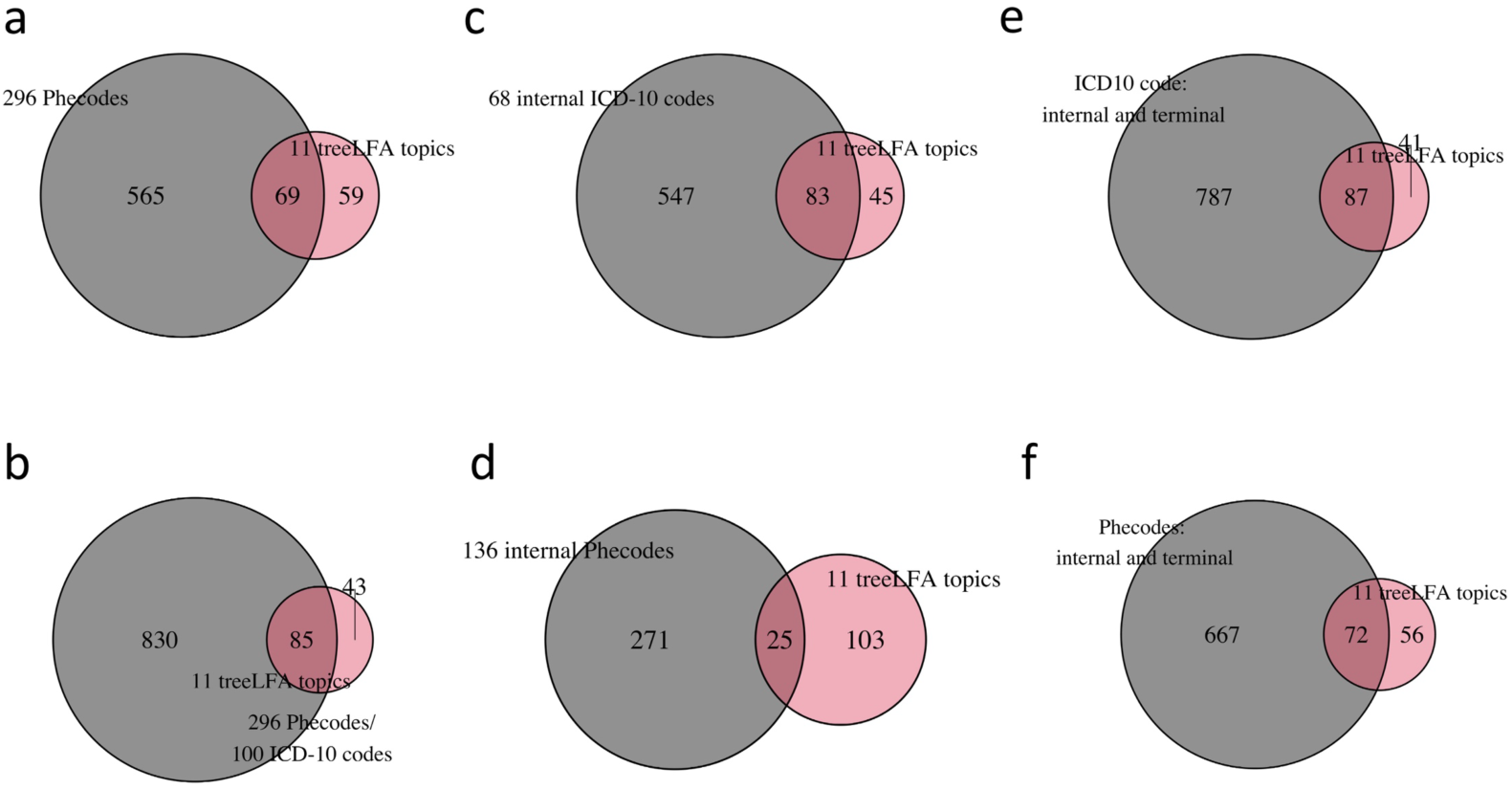
Overlap of significant loci found by GWAS for different traits. a, The total numbers of loci associated with any of the 296 Phecodes mapped from the 100 ICD-10 codes and any of the 11 treeLFA topics, and their overlap. b, The total numbers of loci associated with any of the 296 Phecodes or the 100 ICD-10 codes and any of the 11 treeLFA topics, and their overlap. c, The total numbers of loci associated with any of the 68 internal ICD-10 codes and any of the 11 treeLFA topics, and their overlap. d, The total numbers of loci associated with any of the 136 internal Phecodes and any of the 11 treeLFA topics, and their overlap. e, The total numbers of loci associated with any of the internal or terminal ICD-10 codes and any of the 11 treeLFA topics, and their overlap. f, The total numbers of loci associated with any of the internal or terminal Phecodes and any of the 11 treeLFA topics, and their overlap.

**Supplementary Figure 4.**
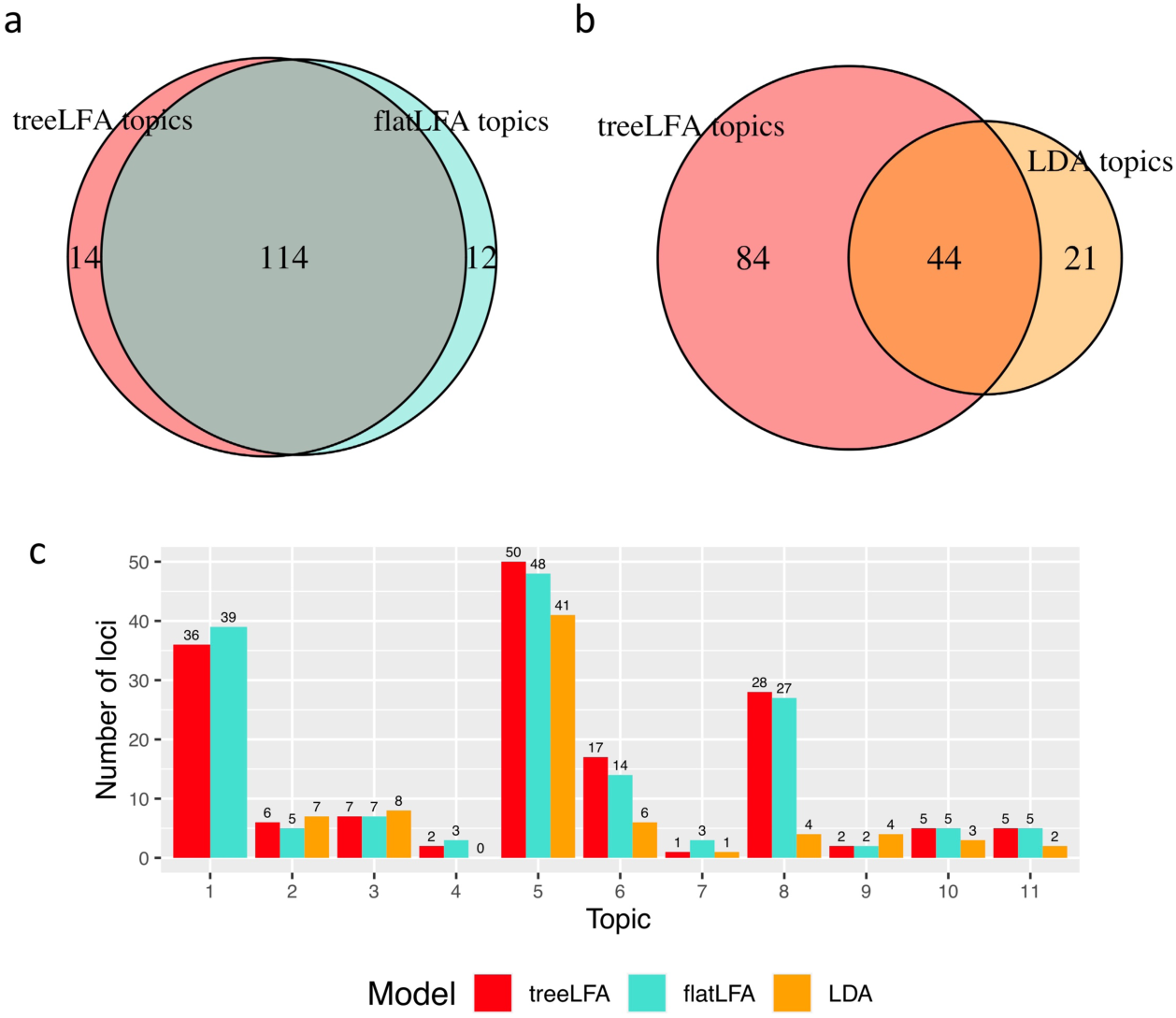
Comparison of the topic-GWAS results for three topic models. a, The total numbers of loci associated with any of the 11 topics inferred by treeLFA and flatLFA, and their overlap. b, The total numbers of loci associated with any of the 11 topics inferred by treeLFA and any of the 10 topics inferred by LDA, and their overlap. c, The numbers of loci associated with each of the 11 topics inferred by treeLFA and flatLFA, and each of the 10 topics inferred by LDA. The first topic is the empty topic, and is only inferred by treeLFA and flatLFA.

**Supplementary Figure 5.**
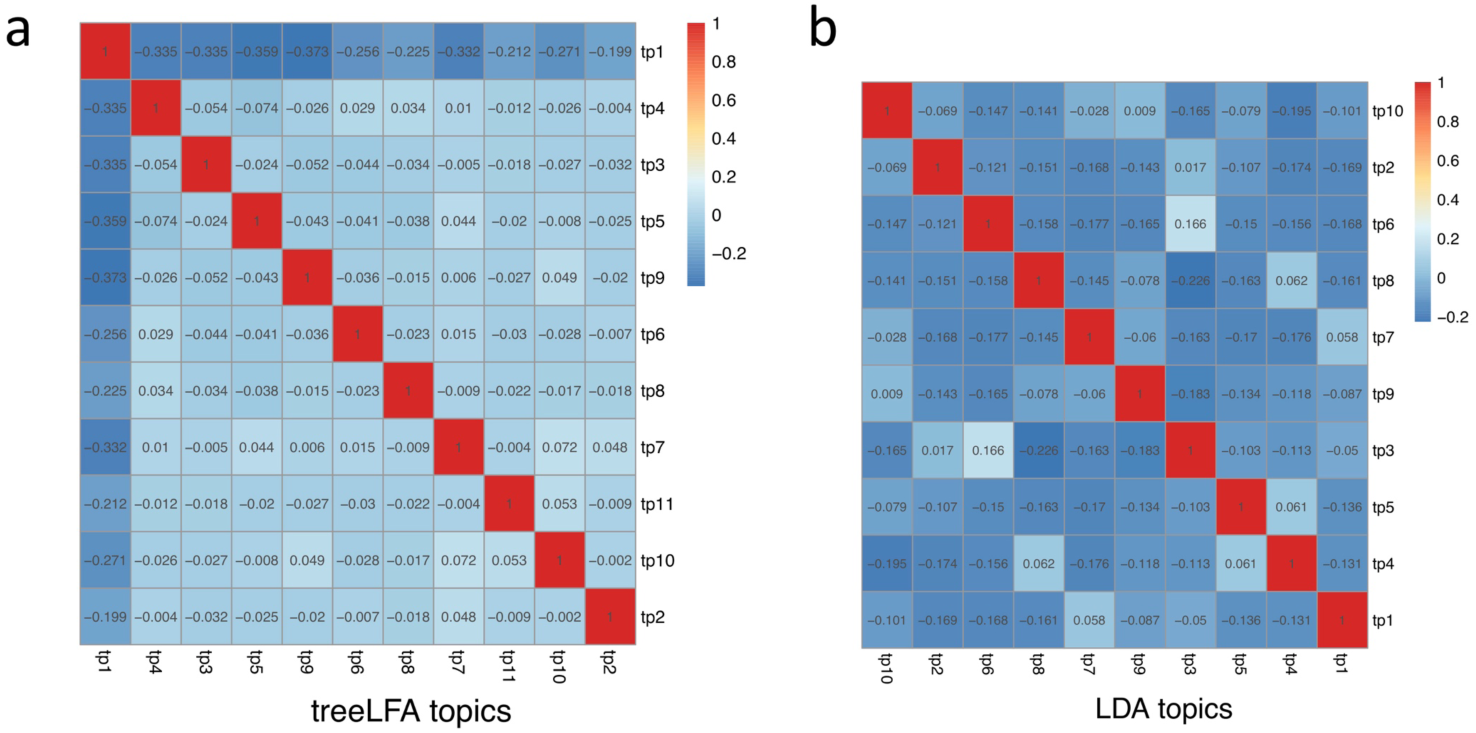
Correlation of topic weights for topics inferred by treeLFA and LDA. a, The correlation matrix for individuals’ weights for the 11 topics inferred by treeLFA. The first topic is the empty topic. b, The correlation matrix for individuals’ weights for the 10 topics inferred by LDA. The matrix uses the same colour scheme as the matrix in panel a.

**Supplementary Figure 6.**
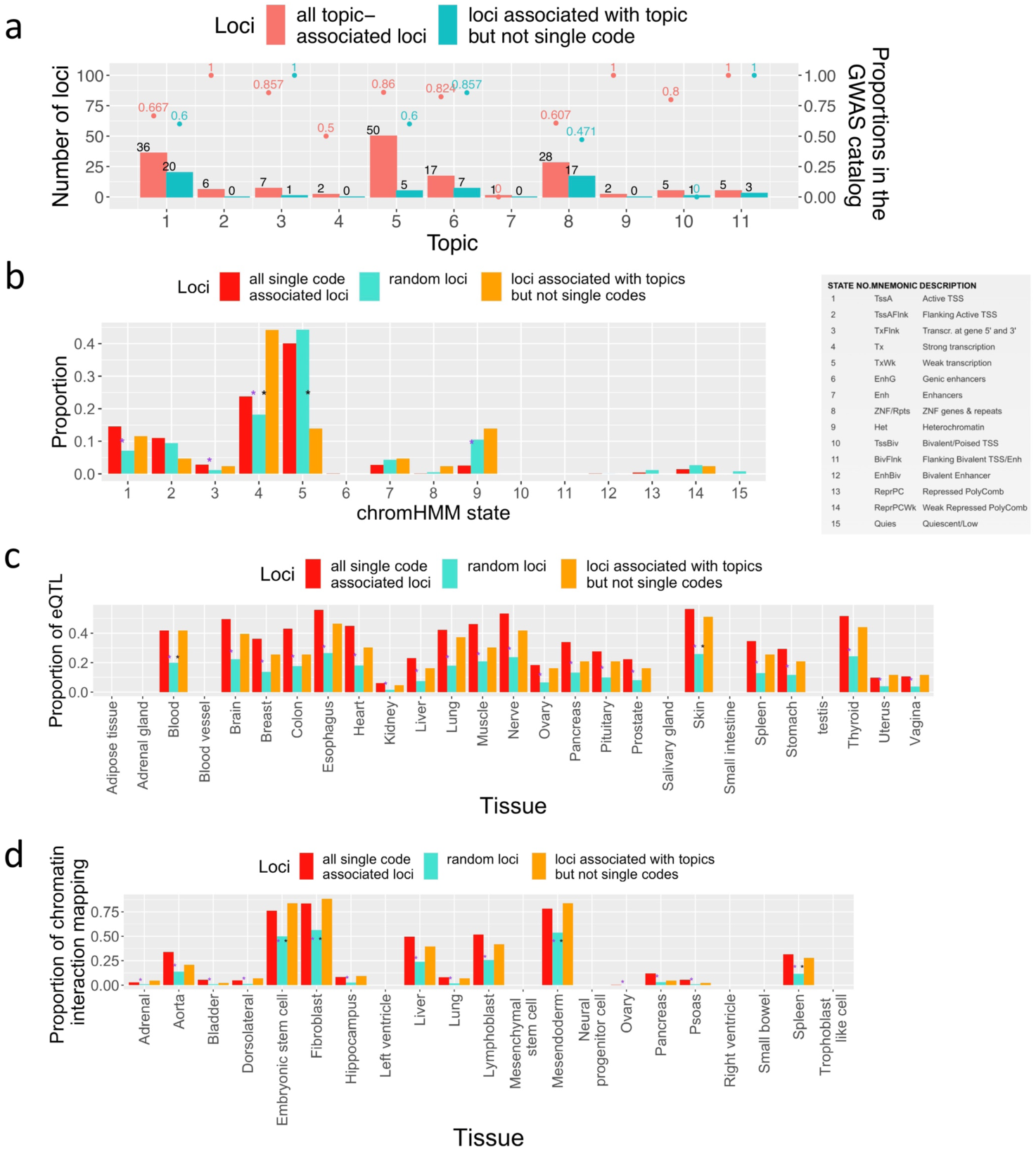
Validation of topic associated loci. a, The proportions of topic-associated loci recorded in the GWAS catalogue. Red bars show the results for all topic-associated loci, and green bars show the results for loci that are associated with topics but not any single code. b, Proportions of three groups of SNPs that are in different chromHMM states. Meanings of different chromHMM states are shown in the right table. The first group contains lead SNPs associated with at least one ICD-10 code; the second group contains 10,000 random SNPs whose allele frequency is matched with that of topic-associated lead SNPs; The third group contains lead SNPs associated with at least one of the 11 topics but not any single code. The proportions of the first and third groups are compared with the second group respectively, and significant differences in proportions (two-proportion Z-test, adjusted P-value<0.05, Bonferroni correction) are marked with asterisks between the corresponding bars (purple asterisks between red and green bars mean significant differences between the first and second groups, black asterisks between blue and green bars mean significant differences between the third and second groups). The numeric results are in Supplementary Table 26. c, Proportions of the three groups of SNPs in panel b that are eQTL in different tissues. The numeric results are in Supplementary Table 27. d, Proportions of the three groups of SNPs in panel b that have chromatin interaction with genes in different tissues. The numeric results are in Supplementary Table 28.

**Supplementary Figure 7.**
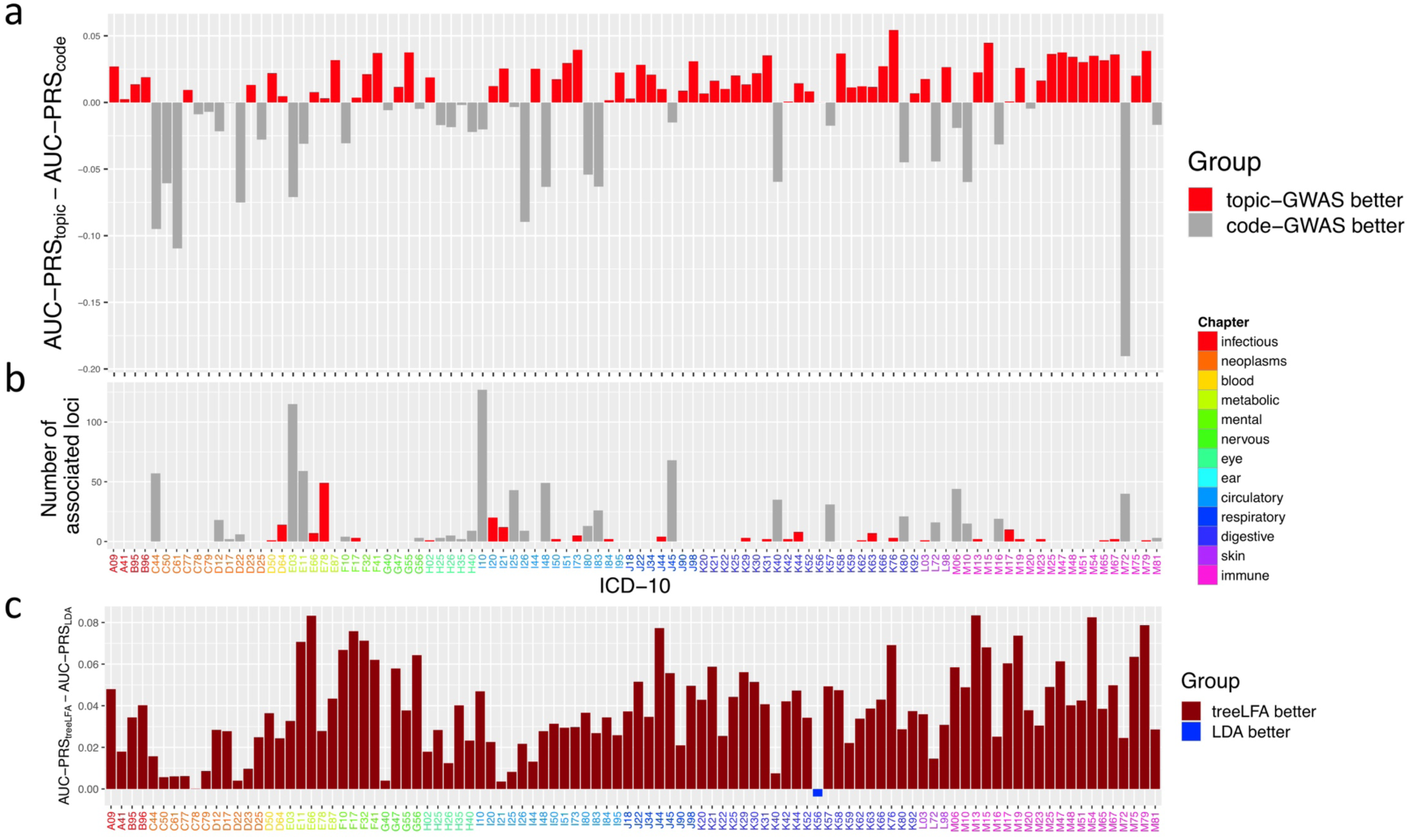
PRS for ICD-10 codes based on topic-GWAS results. a, Comparison of the AUC of two types of PRS for the 100 ICD-10 codes on the test data. One type of PRS is constructed using the topic-GWAS result for treeLFA, and the other type of PRS is constructed using single code GWAS result. Bars are colored according to the relative performance of the two types of PRS. The numeric results are in Supplementary Table 10. b, The numbers of loci associated with each of the 100 ICD-10 codes. Bars are colored the same way as in panel a. Codes are colored according to the ICD-10 chapters they belong to. The numeric results are in Supplementary Table 29. c, Comparison of the AUC of PRS constructed using the topic-GWAS results for treeLFA and LDA. Bars are colored according to the relative performance of the two types of PRS. The numeric results are in Supplementary Table 30.

**Supplementary Figure 8.**
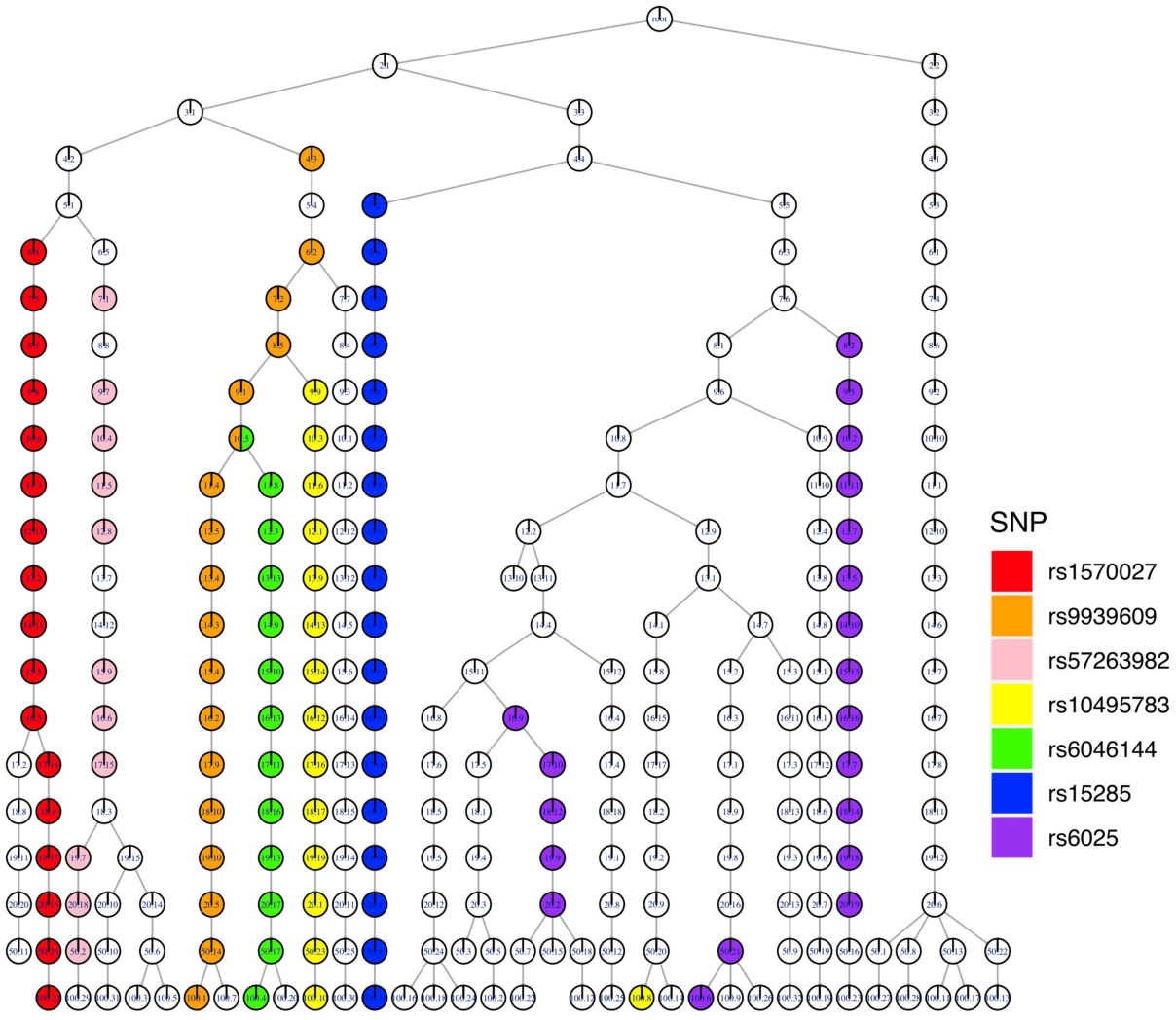
Association of SNPs and topics from different treeLFA models. The associations of a few example SNPs and topics inferred by different treeLFA models are visualised on the tree structure of topics. The tree structure of topics is the same as the one in Figure 5b. Topics significantly associated with 7 different SNPs are highlighted with different colours on the tree structure. The numeric results are in Supplementary Table 31.

**Supplementary Figure 9.**
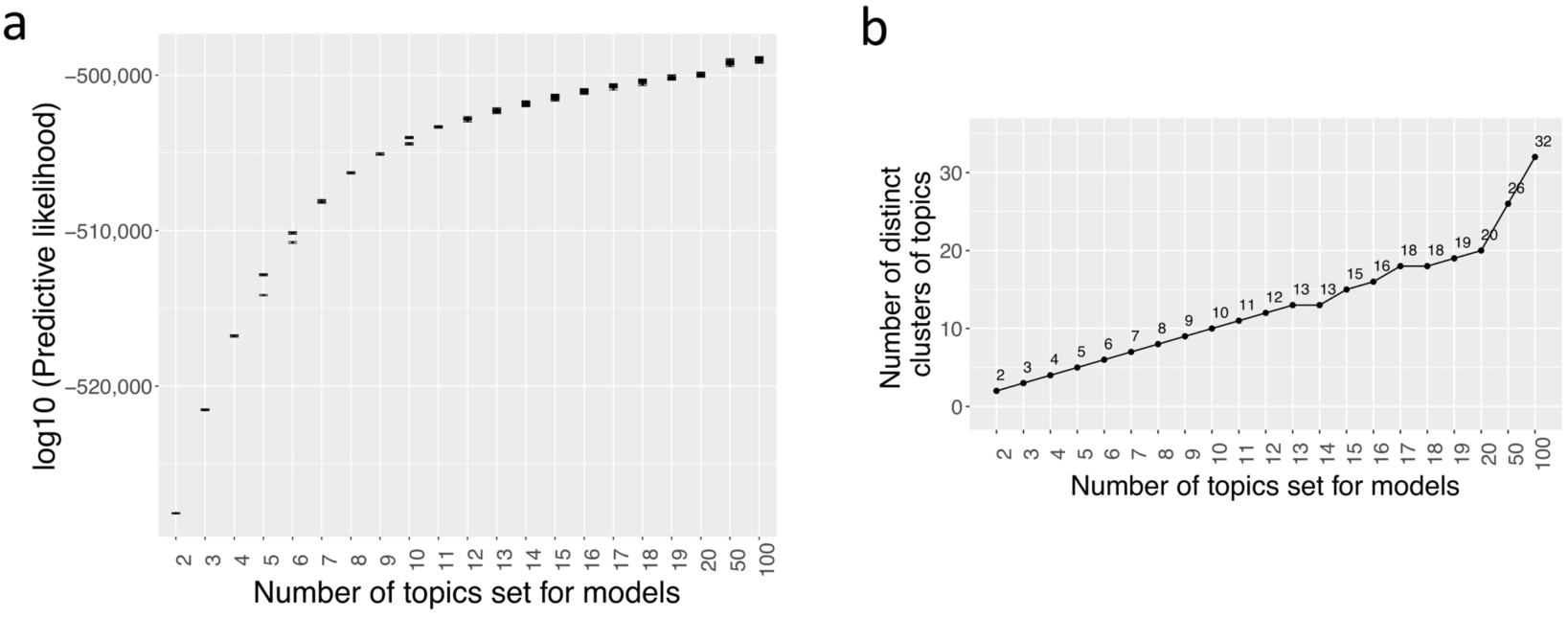
Summary statistics for models with different numbers of topics. a, The predictive likelihood on the test data for the ten Gibbs chains for treeLFA models set with different numbers of topics. For each chain, the standard deviation of the likelihood calculated using different posterior samples of topics were shown. The numeric results are in Supplementary Table 32. b, Numbers of distinct topics remained after clustering for treeLFA models set with different numbers of topics.

**Supplementary figure 10.**
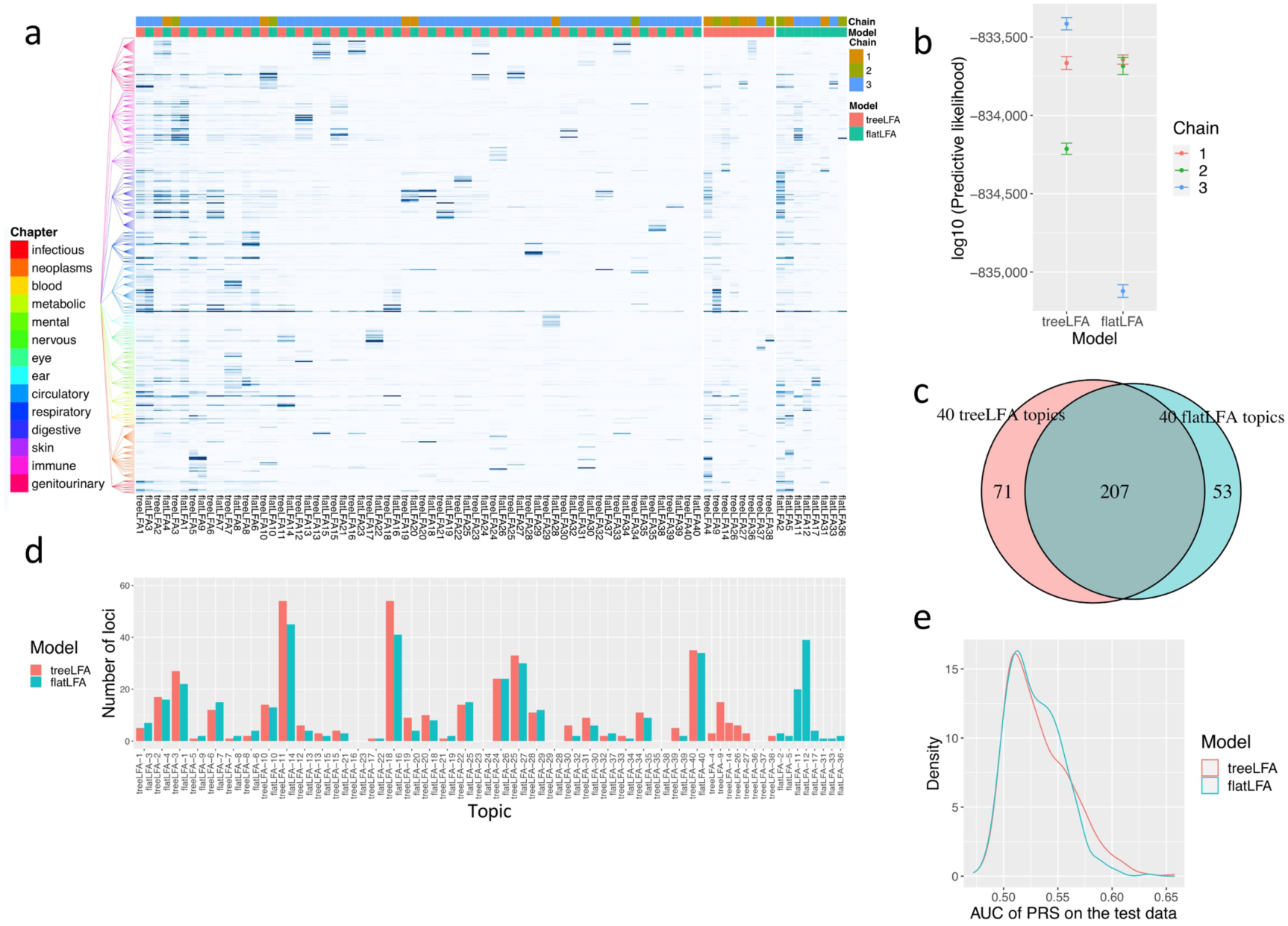
Comparison of the inference and topic-GWAS results for treeLFA and flatLFA on the top-436 UKB dataset. a, 40 topics inferred by treeLFA and flatLFA models set with 100 topics. The same topics inferred by treeLFA and flatLFA are placed next to each other. Topics inferred by both models are shown first, followed by topics inferred by only one model. For each model, the inferred topics are numbered according to their density. The tree structure of the 436 ICD-10 codes is shown to the left of the heatmap, and codes from different ICD-10 chapters are colored differently. For each topic, the number of chains that inferred it is shown with the colour bar on top of the heatmap. The numeric results are in Supplementary Table 33. b, The predictive likelihood on the test data for the three treeLFA and flatLFA chains. The calculation of predictive likelihood was repeated ten times for each chain to get the standard deviation. The numeric results are in Supplementary Table 34. c, The total numbers of loci associated with any of the topics inferred by treeLFA and flatLFA, and their overlap. d, The numbers of loci associated with each of the treeLFA and flatLDA topics. Topics have the same order as those in panel a. The numeric results are in Supplementary Table 35. e, Density plots for the AUC of PRS for the 436 ICD-10 codes on the test data based on the topic- GWAS results for treeLFA and flatLFA. The numeric results are in Supplementary Table 36.

**Supplementary figure 11.**
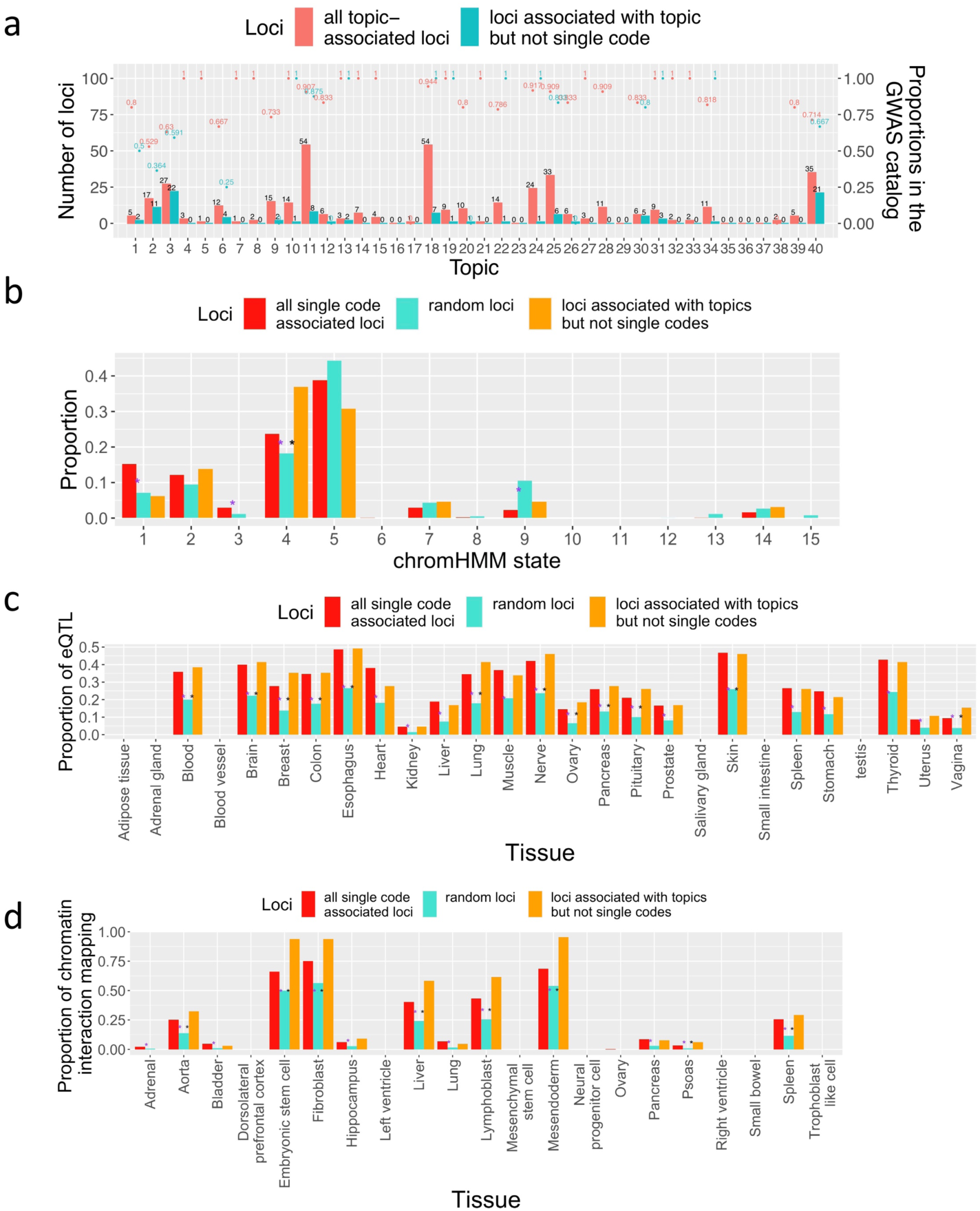
Validation of topic associated loci for the top-436 UKB dataset. All settings are the same as those in Supplementary Figure 6. a, The proportions of topic-associated loci recorded in the GWAS catalogue. b, Proportions of the three groups of SNPs that are in different chromHMM states. The numeric results are in Supplementary Table 37. c, Proportions of the three groups of SNPs in panel b that are eQTL in different tissues. The numeric results are in Supplementary Table 38. d, Proportions of the three groups of SNPs in panel b that have chromatin interaction with genes in different tissues. The numeric results are in Supplementary Table 39.

### Supplementary Tables

**Supplementary Table 1. Parameter setting for all simulated datasets.** 8 groups of datasets are simulated using different combinations of three parameters: the concentration parameter (*α*) of the Dirichlet prior for topic weight variable *θ*, the number of individuals in the training dataset (D), and the tree structure of diseases. Correct tree structure means that the topics used for simulation are likely to be constructed by running a Markov process with small *ρ*_01_ and large *ρ*_11_ (see Methods) on the tree structure.

**Supplementary Table 2. Comparison of 3 topic models on the 4 groups of datasets simulated using correct tree structure of diseases.** Each group contains 20 datasets simulated using the same topics and hyperparameters (Supplementary Table 1). Phi_diff_ave: the averaged per disease difference in probability (Phi) between true and inferred topics. I_diff_ave: the averaged per disease difference in the values of indicator variables (I) between true and inferred topics. Predictive likelihood: predictive likelihood on the corresponding test data, which is calculated using the topics inferred from the training data.

**Supplementary Table 3. Tree structure of the top-100 most frequent ICD-10 codes from chapters 1-13 of the ICD-10 coding system in UK Biobank.** In the first column both internal and terminal nodes of the tree structure of the top-100 most frequent ICD-10 codes are listed (ordered according to the layers of nodes on the tree and the names of nodes). In the second column the parent nodes of the nodes in the first column are shown.

**Supplementary Table 4. The 11 topics inferred by treeLFA on the top-100 UKB dataset.** The probability of the 100 ICD-10 codes in the 11 topics inferred by the treeLFA model set with 11 topics on the top-100 UKB dataset is shown.

**Supplementary Table 5. Topic weights for the 11 topics inferred by treeLFA for 2,000 randomly selected individuals in UKB.**

**Supplementary Table 6. Number of loci associated with individuals’ weights for the 11 topics inferred by treeLFA on the top-100 UKB dataset.**

**Supplementary Table 7. Comparison of -log10 (P-value) for lead SNPs of Topic 5 given by single code GWAS for the top five active codes in Topic 5 and topic-GWAS for Topic 5.**

**Supplementary Table 8. Inflation in P-values given by topic-GWAS for the 11 topics inferred by treeLFA on the top-100 UKB dataset.** Both the intercept and *λ*_GC_ are given by LDSC software (see Methods). *λ*_GC_: genomic control inflation factor.

**Supplementary Table 9. Performance of PRS for topics constructed using topic- GWAS results on the test data.** R2: phenotypic variance of topic weights explained by the PRS for topics. P: P-value of model fit on the testing data. Heritability: heritabilities of topic weights calculated using the LD score regression and summary statistics of topic-GWAS. NUM_SNP: numbers of SNPs used by the software “PRSice-2” to construct the PRS for topics. Threshold: threshold for P-values of SNPs used by “PRSice-2” to construct the PRS for topics.

**Supplementary Table 10. Comparison of the performance of two types of PRS for the 100 ICD10 codes.** One type of PRS for ICD-10 codes is constructed using topic-GWAS results (see Methods), and the other type of PRS is constructed using single code GWAS results. AUC: the area under the receiver-operator curve of the PRS on the test data.

**Supplementary Table 11. Topics inferred by treeLFA models set with different numbers of topics on the top-100 UKB dataset.** The probability of the 100 ICD-10 codes in topics inferred by treeLFA models set with different numbers of topics on the top-100 UKB dataset is shown.

**Supplementary Table 12. topic-GWAS results for the example SNP “rs143384” and all topics inferred by all treeLFA models on the top-100 UKB dataset.** Topics are named using the number of topics set for the corresponding treeLFA model, and the index of the inferred topic for that model. For instance, topic “10.10” means the 10th topic inferred by the treeLFA model set with 10 topics. P: the P-value of the SNP given by the linear regression for the topic weight. BETA: estimated effect size of the SNP. SE: standard error of the estimated effect size.

**Supplementary Table 13. Tree structure of the top-436 most frequent ICD-10 codes from chapters 1-14 of the ICD-10 coding system in UKB.** In the first column both internal and terminal nodes of the tree structure of the top-436 most frequent ICD-10 codes are listed (ordered according to the layers of nodes on the tree and the names of nodes). In the second column the parent nodes of the nodes in the first column are shown.

**Supplementary Table 14. The 40 topics inferred by treeLFA on the top-436 UKB dataset.** The probability of the 436 ICD-10 codes in the 40 topics inferred by treeLFA on the top-436 UKB dataset is shown (see Methods).

**Supplementary Table 15.** Numbers of active codes from different ICD10 chapters in the 40 topics inferred by treeLFA on the top-436 UKB dataset (see Methods).

**Supplementary Table 16. Significant SNPs found by topic-GWAS for the 40 treeLFA inferred topics.** CHR: chromosome. BP: physical position (base pair) of SNP. BETA: estimated effect size (regression coefficient). SE: standard error of the estimated effect size. Topic: the index of the inferred topic for the corresponding treeLFA model. Model: the number of topics set for the treeLFA model.

**Supplementary Table 17.** Effect sizes of lead SNPs associated with the 40 treeLFA inferred topics.

**Supplementary Table 18. Numbers of associated loci for the 40 topics inferred by treeLFA on the top-436 UKB dataset.**

**Supplementary Table 19. treeLFA inferred topics with a substantial number of novel loci found by topic-GWAS.** Dominant ICD10 chapter: the ICD-10 chapter which contributes the majority of active codes in the corresponding topic.

**Supplementary Table 20. Comparison of the performance of two types of PRS for the 436 ICD10 codes.** One type of PRS for ICD-10 codes is constructed using topic-GWAS results (see Methods), and the other type of PRS is constructed using single code GWAS results. AUC: the area under the receiver-operator curve of the PRS on the test data.

**Supplementary Table 21. Enriched gene sets in the GWAS catalogue among genes associated with the 40 treeLFA topics.** GeneSet: names of gene sets in the GWAS catalogue. N: the total numbers of genes in the gene sets. n: the numbers of genes in the gene sets that are associated with the treeLFA topics. P-value: P-values of the enrichment analyses. adjusted-P: adjusted P-values of the enrichment analyses. Genes: genes in the enriched gene sets that are associated with the topics. This table is the output of the “SNP- to-gene” function of the software “FUMA”

**Supplementary Table 22. The correspondence between treeLFA inferred topics and loci level genetically interpretable multimorbidity networks found by the study “A global overview of genetically interpretable multimorbidities among common diseases in the UK Biobank”.** The information of the 9 loci level genetically interpretable multimorbidity networks found by a previous study using the diagnostic data in UKB is shown, together with their corresponding topics inferred by treeLFA on the top-436 UKB dataset. The first 5 columns come from the supplementary results of the paper “A global overview of genetically interpretable multimorbidities among common diseases in the UK Biobank”.

**Supplementary Table 23. Comparison of 3 topic models on the 4 groups of datasets simulated using wrong tree structure of diseases. Each group contains 20 datasets simulated using the same topics and hyperparameters (Supplementary Table 1).** Phi_diff_ave: the averaged per disease difference in probability (Phi) between true and inferred topics. I_diff_ave: the averaged per disease difference in the values of indicator variables (I) between true and inferred topics. Predictive likelihood: predictive likelihood on the corresponding test data, which is calculated using the topics inferred from the training data.

**Supplementary Table 24. The 11 topics inferred by flatLFA on the top100 UKB dataset.** The probability of the 100 ICD-10 codes in the 11 topics inferred by the flatLFA model set with 11 topics on the top-100 UKB dataset is shown.

**Supplementary Table 25. The 10 topics inferred by LDA on the top100 UKB dataset.** The probability of the 100 ICD-10 codes in the 10 topics inferred by the LDA model set with 10 topics on the top-100 UKB dataset is shown.

**Supplementary Table 26. Proportions of SNPs in different chromHMM states for the 3 groups of SNPs on the top-100 UKB dataset.** P(random/single code): P-values for the comparison of proportions for random/single code-associated SNPs. P(random/topic): P- values for the comparison of proportions for random/topic-associated SNPs.

**Supplementary Table 27. Proportions of SNPs that are eQTL in different tissues for the 3 groups of SNPs on the top-100 UKB dataset.** P(random/single code): P-values for the comparison of proportions for random/single code-associated SNPs. P(random/topic): P- values for the comparison of proportions for random/topic-associated SNPs.

**Supplementary Table 28. Proportions of SNPs with chromatin mapping in different tissues for the 3 groups of SNPs on the top-100 UKB dataset.** P(random/single code): P- values for the comparison of proportions for random/single code-associated SNPs.

P(random/topic): P-values for the comparison of proportions for random/topic-associated SNPs.

**Supplementary Table 29. Numbers of significant loci given by single code GWAS for the 100 ICD10 codes.**

**Supplementary Table 30. Comparison of the performance of PRS for the 100 ICD-10 codes based on topic-GWAS results for treeLFA/LDA inferred topics on the top-100 UKB dataset.** AUC: the area under the receiver-operator curve of the PRS on the test data.

**Supplementary Table 31. topic-GWAS results for 7 example SNPs and all topics inferred by treeLFA models set with different numbers of topics on the top-100 UKB dataset. P-values given by topic-GWAS for the examples SNPs are shown.** Topics (in the first column) are named using the number of topics set for the corresponding treeLFA model, and the index of the inferred topic for that model. For instance, topic “10.10” means the 10th topic inferred by the treeLFA model set with 10 topics.

**Supplementary Table 32. Predictive likelihood for treeLFA models set with different numbers of topics on the top-100 UKB dataset.** For each treeLFA model, 10 Gibbs chains are trained, and 50 posterior samples of topics are collected from each chain. The predictive likelihood on the test data is calculated using each posterior sample of topics, and standard error of predictive likelihood is calculated for each chain.

**Supplementary Table 33. The 40 topics inferred by treeLFA and flatLFA on the top-436 UKB dataset.** The probability of the 436 ICD-10 codes in the 40 topics inferred by treeLFA and flatLFA is shown.

**Supplementary Table 34. Predictive likelihood on the test data for three treeLFA and flatLFA chains on the top-436 dataset.** For each chain, the predictive likelihood is calculated 10 times.

**Supplementary Table 35. Numbers of associated loci for the 40 treeLFA and flatLFA topics on the top-436 UKB dataset. Topics are named according to their order in Supplementary Table 33.**

**Supplementary Table 36. Comparison of the performance of PRS for the 436 ICD10 codes based on topic-GWAS results for treeLFA and flatLFA topics. AUC: the area under the receiver-operator curve of the PRS on the test data.**

**Supplementary Table 37. Proportions of SNPs in different chromHMM states for the 3 groups of SNPs on the top-436 UKB dataset.** P(random/single code): P-values for the comparison of proportions for random/single code-associated SNPs. P(random/topic): P- values for the comparison of proportions for random/topic-associated SNPs.

**Supplementary Table 38. Proportions of SNPs that are eQTL in different tissues for the 3 groups of SNPs on the top-436 UKB dataset.** P(random/single code): P-values for the comparison of proportions for random/single code-associated SNPs. P(random/topic): P- values for the comparison of proportions for random/topic-associated SNPs.

**Supplementary Table 39. Proportions of SNPs with chromatin mapping in different tissues for the 3 groups of SNPs on the top-436 UKB dataset. P(random/single code): P- values for the comparison of proportions for random/single code-associated SNPs.** P(random/topic): P-values for the comparison of proportions for random/topic-associated SNPs.

