## Supplementary material for "Topic modelling with ICD10-informed priors identifies novel genetic loci associated with multimorbidities in UK Biobank": Analytic note

Yidong Zhang

December 11, 2022

#### 1 Model overview

treeLFA is the the abbreviation for "latent factor allocation with a tree structured prior for topics". It is a topic model specifically designed for binary input data, which is based on the Bayesian mean-parameterized binary non-negative matrix factorization [1], and also incorporates an informative prior for topics constructed on a tree structure of individual words. It retains the basic "document-topic-word" configuration of topic models, but these components are assigned with new meanings to analyse the diagnostic data in biobanks. For treeLFA, each individual in the biobank is viewed as a document, and each disease a word. Topics of diseases capture constellations of diseases that frequently co-occur on the same individuals.

The input for treeLFA is a $D \times S$ binary matrix ($W$), where each row corresponds to an individual ($d$), and each column a disease code ($s$). $W_{ds}$ in the input matrix records if disease code $s$ is diagnosed for individual $d$. The output of treeLFA includes two matrices: the topic matrix $\phi$ ($T \times S$, $\phi_{ts}$ is the probability of disease code $s$ in topic $t$), and the topic weight matrix $\theta$ ($D \times T$, $\theta_{dt}$ is the weight of individual $d$ for topic $t$).

treeLFA is generalised from Bayesian mean-parameterized binary non-negative matrix factorization (BNMF), which is fundamentally different from the more commonly used topic model Latent Dirichlet Allocation (LDA) [2]. treeLFA models the presence and absence of all disease codes for all individuals with Bernoulli distributions, while LDA models the disease codes that are diagnosed (present) for individuals with Multinomial distributions. For treeLFA, the loading of a disease code in a topic is its Bernoulli probability, and the Bernoulli probability of this disease code for an individual is a mixture of the Bernoulli probability for this code in all topics, with the mixing coefficients specified by the individual's topic weights. As a whole, the matrix of Bernoulli probability for all disease variables ($D \times S$ variables in total) is factorised into the topic weight matrix and the topic matrix that are mentioned above. To differentiate from LDA, we name our model latent factor allocation (LFA). In the next sections, we will introduce the generative process for treeLFA, starting from the generation of topics using a hierarchical prior.

#### 2 The hierarchical prior for topics

In each topic, only a fraction of all disease codes are active, which means they have a relatively large Bernoulli probability in the topic. As a result, they will be likely to co-occur on the same individual. The remaining disease codes are inactive, meaning they have near-zero probability. Different Beta priors ($Beta(a_0^0, a_0^1)$ and $Beta(a_1^0, a_1^1)$) are put on $\phi_{ts}$ for inactive and active disease codes.

To introduce correlation in active diseases within each topic, we used an tree-structured prior. For disease code $s$ in topic $t$ a binary indicator variable $I_{ts}$ is introduced to encode whether it is active in the topic (1 for active state, 0 for inactive state). The indicator variables for all disease codes in a topic are generated with a Markov process on the hierarchical/tree structure of disease codes specified by a medical ontology (such as the ICD-10 coding system). As is shown in Figure 1, each leaf node of the tree corresponds to the indicator variable of a disease code in a topic, while internal nodes on the tree correspond to different categories of diseases specified by the medical ontology (here we assume that they can also be diagnosed for individuals). $I$ for all disease codes in a topic are sampled from the inactive root node (does not correspond to any disease code) to all leaf nodes, using a Markov process that has two states, active and inactive. $\rho_{01}$ and $\rho_{11}$ are the transition probability of the Markov process, corresponding to the probability of transitioning from inactive to active state and from active to active state, respectively.

The sparsity of a topic (the total number of active codes in a topic) can be controlled by tuning $\rho_{01}$ and $\rho_{11}$ of the Markov process, as what we can do with LDA by tuning the concentration parameter of the Dirichlet prior for topics. Furthermore, we can also control the distribution of active codes in a topic by tuning the transition probability. With a large $\rho_{11}$, most child codes of an active parent code will also be active. As a result, active codes will be concentrated on the same subtree. On the other hand, if $\rho_{11}$ is small, active codes will be scattered across the whole tree.

#### 3 Generative model specification

The graphical representation of the model is shown in Figure 2A. To generate a topic, $I$ for all disease codes in this topic are firstly generated using a Markov process on the tree structure of disease codes (discussed in the previous section). Next, conditioned on these indicator variables, the probability $\phi$ for all disease codes in the topic are sampled from the corresponding Beta distributions for active and inactive codes. We then generate the topic weight vector $\theta_d$ for individual $d$ using a Dirichlet distribution, and for him the topic assignment variables $Z_d$ for all disease codes using the categorical distribution parameterized by $\theta_d$. These two steps are the same as the corresponding steps in the generative process for LDA (Figure 2B). In the last step, disease variable $W_{ds}$ is sampled from the Bernoulli distribution with probability $\phi_{Z_{ds},s}$ (this is the probability of

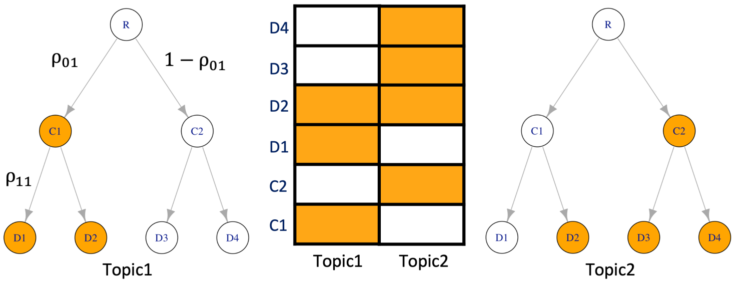


Figure 1: Schematic for the Markov process on the tree structure of disease codes. Values of the indicator variables for six disease codes in two topics are shown in the middle of the figure. Each cell corresponds to the indicator variable of a disease code in a topic, with white color representing inactive code, and orange representing active code. All indicator variables for a topic are generated by running a Markov process on the tree structure of disease codes. Each node (except for the root node) on the tree corresponds to a disease code in a topic. The top node on the tree labelled with $R$ is the single root node, while $C$ represents categories of diseases (internal nodes on the second layer of the tree), and $D$ represents individual diseases (terminal nodes on the third layer of the tree). $\rho_{01}$ and $\rho_{11}$ are the two transition probabilities of this Markov process.

disease code $s$ in the topic specified by topic assignment variable $Z_{ds}$). Notations for treeLFA are listed in Table 1.

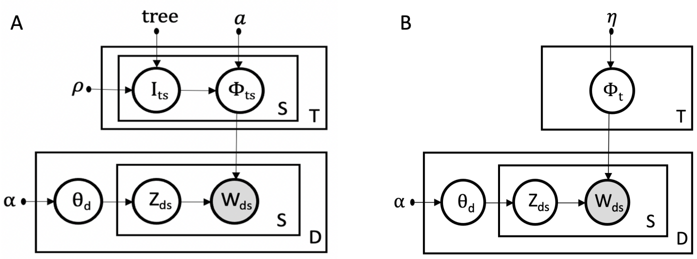


Figure 2: The probabilistic graphical model for treeLFA and LDA. A, The probabilistic graphical model for treeLFA. Each topic is a probability vector $(\phi_{t1}...\phi_{tS})$ for the Bernoulli distributions for $S$ disease codes, and each probability variable $\phi_{ts}$ has a corresponding indicator variable $I_{ts}$, denoting if disease code $s$ is active in topic $t$. Different Beta priors are put on $\phi$ of active and inactive disease codes. $I$ for all disease codes in a topic are generated using a Markov process on the tree structure with transition probability vector $\rho$. The generative steps for the remaining hidden variables are the same as those for LDA. B, The graphical model for LDA.

The full treeLFA model is specified as below:

$$
\begin{aligned}
&P(W, \theta, Z, \phi, I \mid \rho, \alpha, a, tree) \\
=&P(I \mid tree, \rho) \cdot P(\phi \mid I, a) \cdot P(\theta \mid \alpha) \cdot P(Z \mid \theta) \cdot P(W \mid \phi, Z)
\end{aligned} \tag{1}
$$

$$
\begin{aligned}
&P(I \mid tree, \rho) = \prod_{t=1}^{T} \prod_{s=1}^{S} P(I_{ts} \mid I_{ts}^{pa}, \rho) \\
&P(\phi \mid I, a) = \prod_{t=1}^{T} \prod_{s=1}^{S} P(\phi_{ts} \mid I_{ts}, a) \\
&P(\theta \mid \alpha) = \prod_{d=1}^{D} P(\theta_d \mid \alpha) \\
&P(Z \mid \theta) = \prod_{d=1}^{D} \{ \prod_{s=1}^{S} P(Z_{ds} \mid \theta_d) \} \\
&P(W \mid \phi, Z) = \prod_{d=1}^{D} \prod_{s=1}^{S} P(W_{ds} \mid \phi_{Z_{ds}, s}) \\
&P(\theta_d \mid \alpha) \sim Dirichlet(\alpha) \\
&P(Z_{ds} \mid \theta_d) \sim Categorical(\theta_d) \\
&P(I_{ts} \mid I_{ts}^{pa} = 0, \rho) \sim Bernoulli(\rho_{01}) \\
&P(I_{ts} \mid I_{ts}^{pa} = 1, \rho) \sim Bernoulli(\rho_{11}) \\
&P(\phi_{ts} \mid I_{ts} = 0, a) \sim Beta(a_0^0, a_0^1) \\
&P(\phi_{ts} \mid I_{ts} = 1, a) \sim Beta(a_1^0, a_1^1) \\
&P(W_{ds} \mid \phi_{z_{ds}, s}) \sim Bernoulli(\phi_{z_{ds}, s})
\end{aligned}
$$

| Notation | Explanation |
|---|---|
| $D$ | total number of individuals in the training dataset |
| $S$ | total number of disease codes |
| $T$ | total number of topics |
| $d$ | individual $d$ |
| $s$ | disease code $s$ |
| $t$ | topic $t$ |
| $Z_{ds}$ | topic assignment variable for disease code $s$ for individual $d$ |
| $\phi_{ts}$ | probability variable for disease code $s$ in topic $t$ |
| $I_{ts}$ | indicator variable for disease code $s$ in topic $t$ |
| $I_{ts}^{pa}$ | indicator variable for the parent disease code of disease code $s$ in topic $t$ |
| $I_{ts}^{ch}$ | indicator variables for all children disease codes of disease code $s$ in topic $t$ |
| $\theta_d$ | topic weight vector for individual $d$ |
| $W_{ds}$ | disease variable for disease code $s$ for individual $d$ |
| $tree$ | the fixed tree structure of disease codes specified by a disease classification system |
| $\rho_{01}$ | the probability of transitioning from an inactive code to an active code for the Markov process on the tree |
| $\rho_{11}$ | the probability of transitioning from an active code to an active code for the Markov process on the tree |
| $\alpha$ | the parameter vector for the Dirichlet prior for $\theta$ |
| $a_0^0, a_0^1$ | parameters of the Beta prior for $\phi$ of inactive codes |
| $a_1^0, a_1^1$ | parameters of the Beta prior for $\phi$ of active codes |

Table 1: Notations for "treeLFA"

#### 3.1 Inference of treeLFA with Gibbs sampling

We use a partially collapsed Gibbs sampler to estimate the posterior distributions of the latent variables of treeLFA. The collapsed Gibbs sampler performs inference on the marginal distribution of disease incidences, with individuals' topic weight vectors $\theta$ integrated out. The updating equations for each hidden variable are derived as follows:

##### 3.1.1 Topic assignment variable for disease code s for individual d: $Z_{ds}$

$$
\begin{aligned}
P(Z_{ds} = t* \mid Z_d^{\neg s}, \phi, I, W) \propto & \int P(Z_{ds} = t*, Z_d^{\neg s}, \phi, I, W, \theta) d\theta \\
\propto & P(W_{ds} \mid Z_{ds} = t*, \phi) \cdot \int P(\theta_d \mid \alpha) \cdot P(Z_d \mid \theta_d) d\theta_d \\
= & P(W_{ds} \mid Z_{ds} = t*, \phi) \cdot \int [(\frac{\Gamma(\sum_{t=1}^T \alpha_t)}{\prod_{t=1}^T \Gamma(\alpha_t)} \prod_{t=1}^T \theta_{dt}^{\alpha_t - 1}) \\
& \prod_{s=1}^S \theta_{d, Z_{ds}}] d\theta_d \\
= & P(W_{ds} \mid Z_{ds} = t*, \phi) \cdot \int [\frac{\Gamma(\sum_{t=1}^T \alpha_t)}{\prod_{t=1}^T \Gamma(\alpha_t)} \prod_{t=1}^T \theta_{dt}^{\alpha_t - 1} \\
& \prod_{t=1}^T \theta_{dt}^{c_{dt}}] d\theta_d \\
= & P(W_{ds} \mid Z_{ds} = t*, \phi) \cdot \frac{\Gamma(\sum_{t=1}^T \alpha_t)}{\prod_{t=1}^T \Gamma(\alpha_t)} \cdot \frac{\prod_{t=1}^T \Gamma(\alpha_t + c_{dt})}{\Gamma(\sum_{t=1}^T (\alpha_t + c_{dt}))} \\
\propto & P(W_{ds} \mid Z_{ds} = t*, \phi) \cdot \frac{\prod_{t=1}^T \Gamma(\alpha_t + c_{dt})}{\Gamma(\sum_{t=1}^T (\alpha_t + c_{dt}))} \\
= & P(W_{ds} \mid Z_{ds} = t*, \phi) \cdot \frac{\prod_{t \neq t*} \Gamma(\alpha_t + c_{dt}^{\neg s})}{\Gamma([\sum_{t=1}^T (\alpha_t + c_{dt}^{\neg s})] + 1)} \\
& \cdot \Gamma(\alpha_{t*} + c_{dt*}^{\neg s} + 1) \\
= & P(W_{ds} \mid Z_{ds} = t*, \phi) \cdot \frac{\prod_{t=1}^T \Gamma(\alpha_t + c_{dt}^{\neg s})}{\Gamma([\sum_{t=1}^T (\alpha_t + c_{dt}^{\neg s})] + 1)} \\
& \cdot (\alpha_{t*} + c_{dt*}^{\neg s}) \\
\propto & \phi_{t*s}^{W_{ds}} \cdot (1 - \phi_{t*s})^{1 - W_{ds}} \cdot (\alpha_{t*} + c_{dt*}^{\neg s})
\end{aligned}
$$

$$
P(Z_{ds} = t* \mid Z_d^{\neg s}, \phi, I, W) = \frac{\phi_{t*s}^{W_{ds}} \cdot (1 - \phi_{t*s})^{1 - W_{ds}} \cdot (\alpha_{t*} + c_{dt*}^{\neg s})}{\sum_{t=1}^T [\phi_{ts}^{W_{ds}} \cdot (1 - \phi_{ts})^{1 - W_{ds}} \cdot (\alpha_t + c_{dt}^{\neg s})]} \tag{2}
$$

$Z_d^{\neg s}$: topic assignment variables for all disease codes excluding disease code $s$ for individual $d$.
$c_{dt}$: the total number of disease codes assigned with topic t for individual d.
$c_{dt}^{\neg s}$: the total number of disease codes excluding disease $s$ assigned with topic $t$ for individual $d$.

We reduce the computation time by extracting common terms that are used repetitively. To update $Z_{ds}$ we need to calculate the numerator $\phi_{t*s}^{W_{ds}} \cdot (1 - \phi_{t*s})^{1 - W_{ds}} \cdot (\alpha_{t*} + c_{dt*}^{\neg s})$ for all topics, and then normalize them. Depending on whether $W_{ds}$ is zero or one, only one of $\phi_{t*s}^{W_{ds}}$ and $(1 - \phi_{t*s})^{1 - W_{ds}}$ needs to be calculated. Take $\phi_{t*s}^{W_{ds}}$ as an example, when $W_{ds} = 1$ we need to calculate $\phi_{t*s} \cdot (\alpha_{t*} + c_{dt*}^{\neg s})$, which equals $\phi_{t*s} \cdot \alpha_{t*} + \phi_{t*s} \cdot c_{dt*}^{\neg s}$. When sampling $Z_{ds}$, $\phi_{t*s} \cdot \alpha_{t*}$ have fixed values. We can calculate and cache these values for all $t$ and $s$ before sampling $Z$. This simple trick can significantly reduce the time we spend on sampling $Z$.

##### 3.1.2 Topic loading variable for disease code $s$ in topic $t$: $\phi_{ts}$

$$
\begin{aligned}
P(\phi_{ts} \mid \cdot) &\propto P(W \mid Z, \phi_{ts}) \cdot P(\phi_{ts} \mid a, I_{ts}) \\
&\propto [\prod_{d=1}^{D} I(Z_{ds}=t) \cdot P(W_{ds} \mid \phi_{ts})] \cdot P(\phi_{ts} \mid a, I_{ts}) \\
&= [\prod_{d=1}^{D} I(Z_{ds}=t) \cdot (1-\phi_{ts})^{1-W_{ds}} \cdot \phi_{ts}^{W_{ds}}] \cdot Beta(\phi_{ts} \mid a^0_{I_{ts}}, a^1_{I_{ts}}) \\
&\propto \phi_{ts}^{\sum_{d=1,Z_{ds}=t}^{D} W_{ds}} \cdot (1-\phi_{ts})^{\sum_{d=1,Z_{ds}=t}^{D}(1-W_{ds})} \cdot (1-\phi_{ts})^{a^1_{I_{ts}}-1} \cdot (\phi_{ts})^{a^0_{I_{ts}}-1}
\end{aligned}
$$

$$
P(\phi_{ts} \mid \cdot) \sim Beta(a^0_{I_{ts}} + N^1_{st}, a^1_{I_{ts}} + N^0_{st}). \tag{3}
$$

$N^0_{st}$: among individuals who have $W_{ds}$=0, the total number of individuals whose disease variables $s$ are assigned with topic $t$.
$N^1_{st}$: among individuals who have $W_{ds}$=1, the total number of individuals whose disease variables $s$ are assigned with topic $t$.

$$
\begin{aligned}
N^0_{st} &= \sum_{d=1}^{D} I(Z_{ds}=t, W_{ds}=0). \\
N^1_{st} &= \sum_{d=1}^{D} I(Z_{ds}=t, W_{ds}=1).
\end{aligned}
$$

##### 3.1.3 Indicator variable for disease code $s$ in topic $t$: $I_{ts}$

$$
\begin{aligned}
P(I_{ts}=0 \mid \cdot) &\propto P(\phi_{ts} \mid I_{ts}=0) \cdot P(I_{ts}=0 \mid I^{pa}_{ts}) \cdot P(I^{ch}_{ts} \mid I_{ts}=0) \\
&= P(\phi_{ts} \mid I_{ts}=0) \cdot ((1-\rho_{11})^{I^{pa}_{ts}} \cdot (1-\rho_{01})^{1-I^{pa}_{ts}}) \cdot (\rho_{01}^{N^1_{ts,ch}} \cdot (1-\rho_{01})^{N^0_{ts,ch}}) \\
&= Beta(\phi_{ts} \mid a^0_0, a^1_0) \cdot ((1-\rho_{11})^{I^{pa}_{ts}} \cdot (1-\rho_{01})^{1-I^{pa}_{ts}}) \cdot (\rho_{01}^{N^1_{ts,ch}} \cdot (1-\rho_{01})^{N^0_{ts,ch}}).
\end{aligned}
$$

$$
P(I_{ts}=1 \mid \cdot) \propto Beta(\phi_{ts} \mid a^0_1, a^1_1) \cdot (\rho_{11}^{I^{pa}_{ts}} \cdot \rho_{01}^{1-I^{pa}_{ts}}) \cdot (\rho_{11}^{N^1_{ts,ch}} \cdot (1-\rho_{11})^{N^0_{ts,ch}}). \tag{4}
$$

We have omitted the uninformative Bernoulli prior on $I_{ts}$ which is cancelled out when normalising $P(I_{ts}=1 \mid \cdot)$ and $P(I_{ts}=0 \mid \cdot)$ such that they sum to 1.
$I^{pa}_{ts}$: the indicator variable for the parent code of code $s$ in topic $t$.
$I^{ch}_{ts}$: the indicator variables for all the children codes of code $s$ in topic $t$.
$N^1_{ts,ch}$: the total number of indicator variables that equal 1 among all the children codes of code $s$ in topic $t$.
$N^0_{ts,ch}$: the total number of indicator variables that equal 0 among all the children codes of code $s$ in topic $t$.

### 4 Learn hyperparameters of treeLFA

There are three hyperparameters for treeLFA, including vector $\alpha$ that parameterise the Dirichlet prior for topic weight vector $\theta$, the transition probability vector $\rho$ for the Markov process on the tree structure, and vector $a$ that parameterise the Beta priors for the probability variables $\phi$.

#### 4.1 Beta prior for transition probability of the Markov process on the tree structure

The inference of the transition probability vector $\rho$ of the Markov process is integrated into the Gibbs sampling framework for treeLFA by putting Beta priors on them as follows:

$$
\begin{aligned}
P(\rho_{01}) &\sim Beta(b_{00}, b_{01}) \\
P(\rho_{11}) &\sim Beta(b_{10}, b_{11})
\end{aligned}
$$

The updating equations for these transition probability variables are:

$$
\begin{aligned}
P(\rho_{01} \mid I) &\sim Beta(N_1^0 + b_{00}, N_0^0 + b_{01}) \\
P(\rho_{11} \mid I) &\sim Beta(N_1^1 + b_{10}, N_0^1 + b_{11})
\end{aligned}
\tag{5}
$$

$N_1^0$: the total number of active indicator variables in all topics with an inactive parent indicator variable.
$N_1^1$: the total number of active indicator variables in all topics with an active parent indicator variable.
$N_0^0$: the total number of inactive indicator variables in all topics with an inactive parent indicator variable.
$N_0^1$: the total number of inactive indicator variables in all topics with an active parent indicator variables.

#### 4.2 Beta prior for probability of disease codes in topics

We use pre-specified beta distributions as the prior for $\phi$. In Figure 3 we show the histograms of 10,000 samples drawn from Beta(0.3,80) and Beta(2,4), which are the prior distributions we choose for $\phi$ of inactive and active codes in topics used for simulation and inference on the top-100 UKB dataset.

#### 4.3 Optimization of asymmetric Dirichlet prior for topic weights

We choose to estimate hyperparameter $\alpha$ to allow for the flexibility that different topics are assigned to individuals' disease codes with varying probability. For LDA it is common to use a symmetric (in most cases an uninformative) Dirichlet prior [3] parameterized by $\alpha$, which implicitly assumes that there are

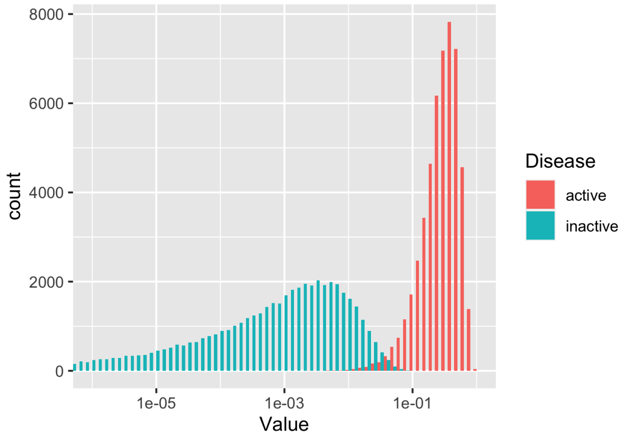


Figure 3: Histogram of samples from the Beta priors for probability of disease codes in topics. Different Beta priors are used for the probability variable $\phi$ for active and inactive codes in topics, such that active codes tend to have large probability, while inactive codes have negligible small probability. 10,000 samples are drawn from Beta(0.3,80) and Beta(2,4) respectively, and their histograms are plotted.

roughly same number of words assigned to each topic across documents. However, the prevalence of diseases in biobanks varies greatly, from one in thousands to more than 0.1 (for example, 28.2 % individuals in UKB are diagnosed with essential hypertension), calling for different topic frequencies. In addition, an empty topic will always be inferred by treeLFA on real-world diagnostic data, and assigned to most disease variables that have value zero. Consequently, the empty topic will have a much larger weight in $\alpha$ than other disease topics, as most individuals only have a few diagnosed diseases. This also requires an asymmetric Dirichlet prior distribution for $\theta$ to be used.

Wallach *et al.* studied the use of complex Dirichlet prior for topic models, and advocated using asymmetric Dirichlet prior for topic weight vector $\theta$ and symmetric Dirichlet prior for topic loading $\phi$ for LDA. He verified with experiments that this strategy of choosing hyperparameters gave the best inference result. Besides, he also suggested the use of an optimization based method to estimate $\alpha$ from the data, since it gave result as good as that can be achieved by using a full Bayesian approach, and was much simpler to implement and time-saving [4].

We use a stochastic EM algorithm, named "Gibbs-EM" to optimise $\alpha$ [5]. The objective function for this method is the marginal likelihood of the data: $P(W \mid \alpha)$, which is maximised with respect to $\alpha$ by alternating between two steps. In the E-step, we fix the value of $\alpha$ and maximise a lower bound of the marginal likelihood by approximating the posterior distribution of hidden variables with $G$ samples given by the Gibbs sampler for treeLFA:

$$
\begin{aligned}
logP(W \mid \alpha) &\geq \sum_{Z} P(Z \mid W, \alpha) \cdot logP(W, Z \mid \alpha) \\
&\approx \frac{1}{G} \sum_{g=1}^{G} logP(W, Z^g \mid \alpha) \\
&= \frac{1}{G} \sum_{g=1}^{G} [\sum_{d=1}^{D} logP(W_d, Z_d^g \mid \alpha)].
\end{aligned} \tag{6}
$$

In the above equation, hidden variables $\phi$, $I$ and $\theta$ are omitted, and only $Z$ is retained, since it is the only hidden variable related to the optimization of $\alpha$ in the M-step.

In the M-step, we use a fixed point method to optimise $\alpha$ such that the expectation calculated in the E-step is maximised. Entries of $\alpha$ are optimised one at a time:

$$
\alpha_t^{new} = \alpha_t \cdot \frac{\sum_g \sum_d \{\Psi(N_{dt}^g + \alpha_t) - \Psi(\alpha_t)\}}{\sum_g \sum_d \{\Psi(S + \sum_k (\alpha_k)) - \Psi(\sum_k (\alpha_k))\}}. \tag{7}
$$

$N_{dt}^g$: the total number of disease variables for individual $d$ that are assigned with topic $t$ in posterior sample $g$;

$\Psi(\cdot)$: the digamma function.

$S$: the total number of disease codes.

### 5 Model selection: predictive likelihood on the testing dataset

We use predictive likelihood on the held out test data to evaluate models with different topic numbers. We use posterior samples of $\phi$ and the optimal $\alpha$ estimated within the training data to compute the predictive likelihood on the test data. With test data W' and one posterior sample of $\phi$, the predictive likelihood on the test data could be expressed as [6]:

$$P(W^{'} \mid W) = \prod_{d=1}^{D} P(W_d^{'} \mid \phi, \alpha)$$

Ideally, calculation of the predictive likelihood requires integrating out all latent variables. Since this is analytically intractable, we use a Monte-Carlo approximation for this integral. For topic weight variable $\theta_d$, we draw P samples from its prior distribution and use them to approximate the integral:

$$\begin{aligned} &\theta_d^p \sim Dirichlet(\alpha) \\ &P(W_d^{'} \mid \phi, \alpha) \approx \frac{1}{P} \sum_{p=1}^{P} P(W_d^{'} \mid \theta_d^p, \phi, \alpha) \end{aligned}$$

Conditioned on one sample of $\theta_d$ for individual $d$, we calculate the likelihood of each disease variable ($W_{ds}$) independently. We do this by summing out $Z_{ds}$ for all topics:

$$\begin{aligned} P(W_d^{'} \mid \theta_d^p, \phi) &= \prod_{s=1}^{S} [\sum_{Z_{ds}=1}^{T} P(W_{ds}^{'}, Z_{ds} \mid \theta_d^p, \phi)] \\ &= \prod_{s=1}^{S} [\sum_{Z_{ds}=1}^{T} P(W_{ds}^{'} \mid \phi_{Z_{ds},s}) \cdot P(Z_{ds} \mid \theta_d^p)] \end{aligned}$$

In summary, the predictive likelihood on the full test data could be expressed as:

$$P(W^{'} \mid W) \approx \prod_{d=1}^{D} \{ \frac{1}{P} \sum_{p=1}^{P} \{ \prod_{s=1}^{S} \{ \sum_{Z_{ds}=1}^{T} [P(W_{ds}^{'} \mid \phi_{Z_{ds},s}) \cdot P(Z_{ds} \mid \theta_d^p)] \} \} \} \tag{8}$$

[5] Thomas Minka. Estimating a dirichlet distribution, 2000.

[6] Andrew Kachites McCallum. Mallet: A machine learning for language toolkit. *http://mallet. cs. umass. edu*, 2002.
